# An improved polygenic score for Parkinson’s disease partly explains variable penetrance of genetic Parkinson’s disease

**DOI:** 10.1101/2025.02.28.25323076

**Authors:** Sebastian Sendel, Zied Landoulsi, Katja Lohmann, Björn-Hergen Laabs, Meike Kasten, Eva-Juliane Vollstedt, Tatiana Usnich, Alexander Balck, Daniela Berg, Dheeraj Reddy Bobbili, Max Borsche, COURAGE-PD Consortium, Andre Franke, Henrike Hanssen, Emadeldin Hassanin, Andrew A. Hicks, Ulrike M. Krämer, Rejko Krüger, Gregor Kuhlenbäumer, Lara M. Lange, Wolfgang Lieb, Brit Mollenhauer, NCER-PD Consortium, Miriam Neis, Peter P. Pramstaller, Jannik Prasuhn, Eva Schaeffer, Manu Sharma, Meike Steinbach, Claudia Trenkwalder, Norbert Brüggemann, Ana Westenberger, Michael Wittig, Inke R. König, Patrick May, Christine Klein, Amke Caliebe

**Affiliations:** Institute of Medical Informatics and Statistics, Christian-Albrechts-University of Kiel, University Hospital Schleswig-Holstein, Campus Kiel, Kiel, Germany; Luxembourg Centre for Systems Biomedicine, University of Luxembourg, Esch-sur-Alzette, Luxembourg; Luxembourg Institute of Health, Strassen, Luxembourg; Institute of Neurogenetics, University of Lübeck, Lübeck, Germany; Institute of Medical Biometry and Statistics, University of Lübeck, Lübeck, Germany; Department of Psychiatry, University Hospital Schleswig-Holstein, Campus Lübeck, Lübeck, Germany; Department of Neurology, University Hospital Schleswig-Holstein, Campus Lübeck, Lübeck, Germany; Department of Neurology, Christian-Albrechts-University of Kiel, University Hospital Schleswig-Holsten, Campus Kiel, Kiel, Germany; Institute of Clinical Molecular Biology, Christian-Albrechts-University of Kiel, Kiel, Germany; Institute for Biomedicine, Eurac Research, Bolzano, Italy; Institute of Medical Psychology, Center of Brain, Behavior and Metabolism, University of Lübeck, Lübeck, Germany; Parkinson Research Clinic, Centre Hospitalier de Luxembourg, Strassen, Luxembourg; Institute of Epidemiology, Christian-Albrechts-University of Kiel, Kiel, Germany; Paracelsus Elena Clinic, Kassel, Germany; Department of Neurology, University Medical Center, Georg August University, Göttingen, Germany; Department of Midwifery Science, University of Lübeck, Lübeck, Germany; Centre for Genetic Epidemiology, Institute for Clinical Epidemiology and Applied Biometry, University of Tübingen, Tübingen, Germany; Department of Neurosurgery, University Medical Center Göttingen, Göttingen, Germany

## Abstract

While genetic causes are identified in up to 15% of all Parkinson’s disease (PD) patients, the remaining idiopathic PD (iPD) patients are attributed to polygenic risk, environmental and lifestyle factors, and interactions thereof. We applied five advanced polygenic score (PGS) tools to data from 1,762 iPD patients and 4,227 healthy controls of European ancestry, resulting in the development of a novel iPD-PGS with significantly improved discriminative performance compared to existing models with an AUC of 0.680 (95% confidence interval (-CI): [0.665, 0.695]). Validation in independent cohorts confirmed its robustness. Notably, patients with early-onset iPD exhibited markedly high PGS values when compared to late-onset iPD patients and healthy controls, underlining the high polygenetic burden in these individuals. We subsequently applied our novel iPD-PGS to carriers of heterozygous *PRKN* variants and *GBA1* coding risk variants. In both cases, our findings suggest that part of the penetrance in these genetic forms of PD can be explained by the same polygenic alterations as observed mitigating iPD. The discriminative potential was greater for *GBA1* than for *PRKN* (*GBA1*: AUC=0.639, 95%-CI=[0.590, 0.687], *PRKN*: AUC=0.594, 95%-CI=[0.501, 0.687]). Our study highlights the potential of advanced PGSs in PD research, particularly for understanding varying penetrance in genetic PD and identifying high-risk individuals.

## Introduction

Parkinson’s disease (PD) is the fastest growing neurological disease with an almost threefold increase in PD prevalence from 1990 to 2021^1–5^ and presents one of the greatest health-related challenges due to the demographic development towards an aging population.

In idiopathic PD (iPD) the exact etiology is unknown and is believed to be influenced by genetic and environmental factors. Also interactions between these components have been identified^6–9^. Its heritability is estimated at around 27%^10^ and genome-wide association studies (GWAS) underlined a highly polygenic nature of the heritability of iPD. This polygenicity can be captured by polygenic scores (PGSs). PGSs can estimate the genetic disease predisposition of each individual by adding the effects of a large number of genome-wide risk variants into a single value. They enable and improve research about the genetic background and overlap of diseases, disease subtypes and symptoms, and gene-environment interactions, as well as the identification of individuals at high risk^11,12^. PGSs are based upon GWAS. The largest European GWAS to date included 56,306 European PD patients and proxy cases and 1,417,791 healthy individuals and identified 90 genome-wide significant independent loci^13^. Recently, an increasing number of GWAS had been performed on non-European or multi-ancestry cohorts^14–18^. Along with performing the aforementioned largest European PD-GWAS, an advanced PGS for PD was also developed in this study. This PD-PGS demonstrates a good discriminative performance between PD patients and healthy controls and was validated in an independent population^6,7,13,19^. It consists of 1805 single nucleotide polymorphisms (SNPs) and was generated by a clumping-and-thresholding method^20,21^. In the past years, various more refined statistical methods based on larger numbers of SNPs for PGS development have become available, raising the possibility of developing an even better discriminating PGS. To our knowledge, no comparison between different PGS methods has been undertaken for iPD yet. All listed iPD-PGSs in the PGS catalogue^22–24^ (www.pgscatalog.org) use fewer than 2000 SNPs, thus potentially missing relevant parts of the polygenic heritability of iPD. Especially, novel promising Bayesian PGS approaches, such as LDPred2^25,26^, PRS-CSx^27,28^, and LDAK^29^, have not yet been applied to their full potential.

Approximately fifteen percent of PD patients have a genetic form of PD (genetic PD) and carry a genetic causative or risk variant (when including *GBA1* coding risk variant carriers)^30,31^. For example, the *SNCA* gene encoding the α-synuclein protein, was the first to be linked to a form of monogenic PD^32,33^. The penetrance of the variants differs considerably from nearly full penetrance (around 100% for homozygous or compound-heterozygous mutation carriers in *PRKN* or *PINK1*) to low penetrance (around 15% for *GBA1* coding risk variant carriers). Up to now, the contribution of the polygenic component of iPD to the penetrance of genetic PD remains uncertain.

In this study, we aimed to to generate a novel PGS with higher discrimination between iPD patients and healthy controls than previous models by comparing five advanced PGS construction tools utilizing a dataset of 1,762 patients and 4,227 controls of European ancestry. The role of eight genes related to genetic PD in this iPD-PGS was a main focus. The performance of the novel iPD on *GBA1* coding risk variant carriers and on *PRKN* heterozygous carriers was especially relevant to the study’s objective. Overall, our study aimed to investigate the potential of advanced PGSs for iPD and genetic PD research. In the light of the PD pandemic, which calls for new measures for early detection, prevention, and disease-modifying therapy strategies, it is especially relevant to identify high-risk individuals. These can then be targeted for primary prevention in combination with environmental and lifestyle factors, e.g., exposure to neurotoxic agents or smoking.

## Results

### Development of an improved iPD-PGS

For this study, we used individual genotyping data from the ProtectMove consortium (www.protect-move.de). After quality control, 5,623,154 SNPs of 6,826 individuals with genetically determined European ancestry were available. Part of the individuals carried variants of genetic PD on eight PD-related genes (*LRRK2*, *SNCA*, *VPS35*, *CHCHD2*, *PRKN*, *PINK1*, PARK7, *GBA1*) and will be termed variant carriers in the following (including variants rated as variants of unknown significance (VUS), risk-variants (on coding regions of *GBA1)*, likely pathogenic and pathogenic variants; heterozygous carriers included as well).

We split our dataset into a late-onset iPD dataset (n=4,227 controls (mean age-at-sampling (AAS)=55 years) ; n=1,762 cases (mean AAS=70 years, mean age-at-onset (AAO)=63 years), both without variant carriers and with an AAO above 40 years for patients), an early-onset iPD dataset (n=66 cases (mean AAS=45 years, mean AAO=35 years), without variant carriers and with an AA0 < 40 years) and a genetic PD dataset (variants carriers: n=436 controls (non-manifesting carriers, mean AAS=53 years), n=335 cases (mean AAS=67 years, mean AAO=60 years) (Fig. 1, Table 1).

**Figure 1:**
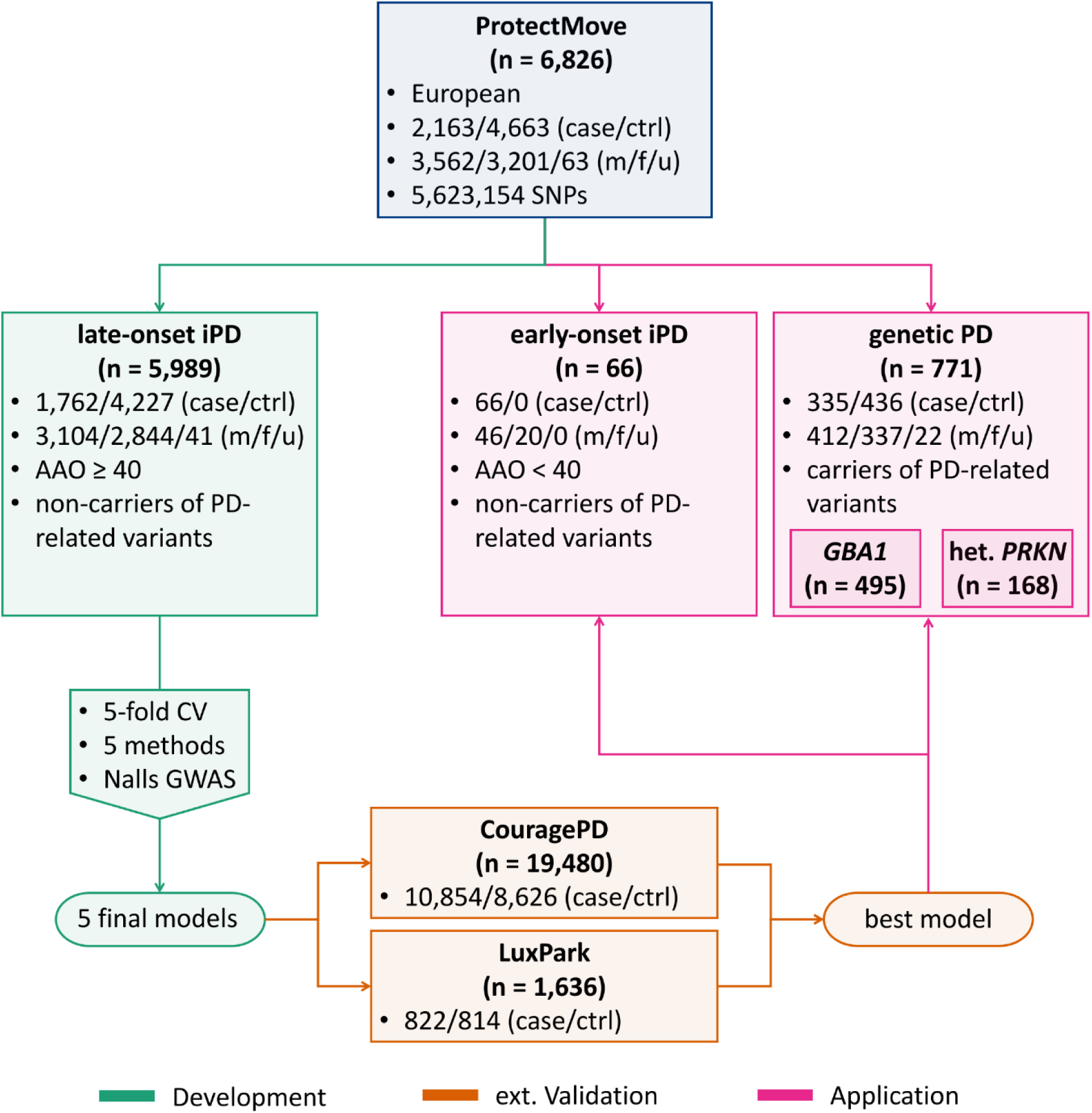
Workflow for PGS model development, validation and application in Parkinson’s disease cohorts. Our ProtectMove cohort (https://protect-move.de/, n = 6,826), comprised European individuals in three datasets: late-onset idiopathic Parkinson’s disease (iPD), early-onset iPD, and variant carriers (PD-related variants (on the genes *LRRK2*, *SNCA*, *VPS35*, *CHCHD2*, *PRKN*, *PINK1*, *PARK7*, *GBA1*) with heterozygous *Parkin* carriers and *GBA1* carriers as largest subgroups, genetic PD). Five tools for generating polygenic scores (PGSs) were applied to the late-onset iPD dataset to derive five final PGS models, based on the GWAS summary statistics from Nalls et al.^13^ and using five-fold cross-validation (CV). The final models were validated in two independent, external cohorts: CouragePD and LuxPark. The best-performing model was identified based on the performance on the late-onset iPD data and the external cohorts (CouragePD, LuxPark). This model was then applied to the genetic PD and early-onset iPD datasets of ProtectMove.

**Table 1:** Characteristics of the ProtectMove datasets used in this study (after quality control)

| Group | N total | N cases | N controls | N cases<br>female | N controls<br>female | AAS cases | AAS controls | AAO cases |
| --- | --- | --- | --- | --- | --- | --- | --- | --- |
| Late-onset iPD | 5,989 | 1,762 | 4,227 | 672 [32] | 2,172 [9] | 70 [63, 76] | 55 [42, 67] | 63 [55, 70] |
| Early-onset iPD | 66 | 66 | 0 | 20 | 0 | 44.5 [39, 54] | - | 35 [29.5, 37] |
| Genetic PD | 771 | 335 | 436 | 126 [11] | 211 [11] | 67 [59, 74] | 53 [43, 64] | 60 [51, 69] |
| **Total** | 6,826 | 2,163 | 4,663 | 818 [43] | 2,383 [20] | 69 [61, 76] | 55 [42, 67] | 62 [54, 70] |

*N*: Total number of samples. *N cases/controls*: Number of cases/controls. *N cases/controls female [NA]*:
Number of female [or unknown sex] cases/controls. *AAS cases/controls*: Median [first and third quartile] for
age-at-sampling of cases/controls, in years. *AAO cases*: Median [first and third quartile] for age-at-onset of
cases, in years. *iPD*: idiopathic Parkinson's disease.

For the development of a new iPD-PGS, the late-onset iPD dataset was used. We applied the following five PGS generating tools, utilizing different statistical approaches, to develop new iPD-PGSs : PRSice-2^20,21^, LDpred2^25,26^ (inf, auto, grid), lassosum2^34,35^, LDAK^29^ (BayesR-SS, Bolt-SS, Ridge-SS), PRS-CSx^27,28^ (Supplementary Methods, Supplementary Table 1). For this, our late-onset iPD dataset was divided into five datasets with equal distribution in case-control status, cohort affiliation, sex and age (AAO, AAS) for five-fold cross validation (CV) (Supplementary Table 2). For each tool, several PGSs were developed and we selected the best-performing one for comparison. Based on all genome-wide variants, the LDpred2 tool demonstrated the best discrimination between PD-patients and healthy controls as measured by the area under the receiver operating characteristic curve (AUC) (Table 2, Fig. 2; for details on the individual methods, see Supplementary Tables 3-7). A mean AUC of 0.679 was achieved on the validation datasets ([minimum, maximum] ([min, max]) across the five datasets: [0.662, 0.691]) and of 0.680 (95% confidence interval (CI): [0.665, 0.695]) on our whole late-onset iPD dataset (n=1,386,717 SNPs in PGS). As comparison, we calculated the AUC for the currently established PGS^13^ (n=1,805 SNPs originally in PGS, 1,097 SNPs available in late-onset iPD dataset) generated with the tool PRSice-2. Here, the resulting AUC was considerably lower, with 0.623 in both the validation and the whole late-onset iPD dataset ([min, max]: validation [0.606, 0.646], whole late-onset iPD 95%-CI: [0.607, 0.638]). The tools LDAK, lassosum2, and PRS-CSx showed a slightly worse performance than LDpred2 with AUCs around 0.66-0.67, whereas PRSice-2 dropped considerably in performance (mean AUC validation: 0.637, [min, max]: [0.623, 0.651]; AUC whole iPD: 0.641, 95%-CI: [0.626, 0.656]).

**Table 2:** Performance of constructed iPD-PGSs on the ProtectMove late-onset iPD dataset

| Tool | CV – all variants |  | Whole late-onset iPD dataset |  |  |  |
| --- | --- | --- | --- | --- | --- | --- |
|  | AUC training | AUC validation | AUC all variants | AUC no PD-genes | AUC no PD-genes, with *SNCA* | #SNPs |
|  | [min, max] | [min, max] | [95%-CI] | [95%-CI] | [95%-CI] |  |
| lassosum2 | 0.665 [0.661, 0.670] | 0.665 [0.648, 0.682] | 0.665 [0.650, 0.680] | 0.656 [0.641, 0.671] | 0.666 [0.651, 0.681] | 21,001 |
| LDAK | 0.662 [0.659, 0.664] | 0.662 [0.655, 0.675] | 0.662 [0.647, 0.677] | 0.656 [0.641, 0.672] | 0.663 [0.648, 0.678] | 3,037,714 |
| **LDpred2** | **0.681 [0.677, 0.684]** | **0.679 [0.662, 0.691]** | **0.680 [0.665, 0.695]** | **0.672 [0.658, 0.687]** | **0.682 [0.667, 0.697]** | **1,386,717** |
| PRS-CSx | 0.663 [0.619, 0.677] | 0.667 [0.607, 0.686] | 0.676 [0.662, 0.691] | 0.665 [0.650, 0.680] | 0.676 [0.661, 0.691] | 766,037 |
| PRSice-2 | 0.642 [0.638, 0.646] | 0.637 [0.623, 0.651] | 0.641 [0.626, 0.656] | 0.632 [0.617, 0.647] | 0.640 [0.625, 0.655] | 1,668 |
| Nalls-PGS | 0.623 [0.617, 0.627] | 0.623 [0.606, 0.646] | 0.623 [0.607, 0.638] | not intended | not intended | 1,097 |

*AUC*: area under the receiver operating characteristic curve. [*min, max*]: minimal and maximal AUC across the
five cross-validation sets. [*95%-CI*]: 95% confidence interval of the AUC. *CV – all variants*: cross-validation
results based on all genome-wide variants. *all variants/no PD Genes/no PD genes, with SNCA*: PGS construction
based on all genome-wide variants / excluding variants on PD-related genes / excluding variants on PD-related
genes except for *SNCA*. *#SNPs*: number of SNPs in the final model trained on the whole iPD dataset. *Nalls-PGS*:
PGS developed by Nalls et al.<sup>13</sup>. The best-performing tool is highlighted in bold.

**Figure 2:**
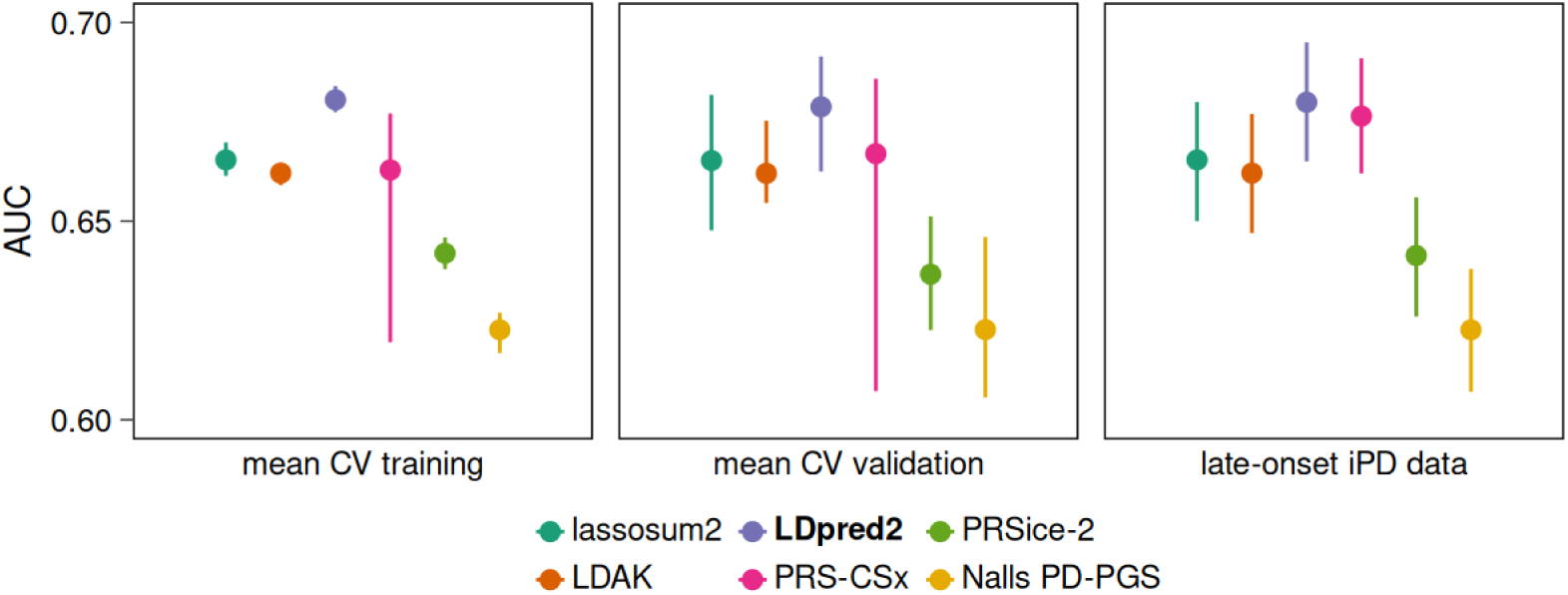
Comparison of performance of constructed iPD-PGSs on the late-onset iPD dataset. Summary of model performance across the five cross-validation (CV) folds and the whole idiopathic late-onset Parkinson’s disease (iPD) dataset, using the area under the receiver operating characteristic curve (AUC) as measure. For the CV approach, the mean AUCs across all five combinations of folds for training and validation were calculated. The error bars indicate the minimum and maximum across the five training or validation datasets. Different colors correspond to the selected tools for polygenic score (PGS) generation with the established PGS by Nalls et al.^13^ as comparison. The overall best-performing and selected final tool (LDpred2) is highlighted in bold.

The best-performing PGS for each tool was applied to two external validation datasets: LuxPark (822 patients and 814 healthy individuals) and CouragePD^36^ (10,854 patients, 8,626 healthy individuals) (Table 3, Fig. 3). Applied to the large CouragePD dataset, the performance of all PGSs was even improved compared to the ProtectMove late-onset iPD dataset. LDAK achieved the highest performance (AUC: 0.717, 95%-CI: [0.709, 0.724]), slightly better than LDpred2 (AUC: 0.715, 95%-CI: [0.708, 0,722]). On the smaller LuxPark dataset, the PGS of lassosum2, LDpred2 and PRS-CSx performed best (AUC: 0.672-0.673), while LDAK (AUC: 0.627) and PRSice-2 (AUC: 0.636) resulted in lower performance. The currently established PGS^13^ showed a solid performance (CouragePD: AUC: 0.663, SNPs available: 1,730; LuxPark: AUC: 0.64, SNPs available: 1,801), even slightly better than PRSice-2 on both datasets.

**Table 3:** Performance of constructed iPD-PGSs on external validation datasets

| Tool |  | AUC LuxPark | #SNPs LuxPark | AUC CouragePD | #SNPs CouragePD |
| --- | --- | --- | --- | --- | --- |
| lassosum2 | f | 0.673 [0.644, 0.702] | 20,967/21,001 | 0.695 [0.688, 0.703] | 20,312/21,001 |
|  | r | 0.659 [0.629, 0.689] | 17,350/17,377 | 0.681 [0.674, 0.689] | 16,816/17,377 |
|  | s | 0.671 [0.641, 0.700] | 20,769/20,803 | 0.691 [0.684, 0.699] | 20,120/20,803 |
| LDAK | f | 0.627 [0.596, 0.657] | 3,032,486/3,037,714 | **0.717 [0.709, 0.724]** | 2,922,433/3,037,714 |
|  | r | 0.619 [0.589, 0.650] | 2,867,958/2,872,825 | **0.711 [0.704, 0.718]** | 2,764,565/2,872,825 |
|  | s | 0.626 [0.595, 0.656] | 2,956,864/2,962,025 | **0.715 [0.708, 0.722]** | 2,848,200/2,962,025 |
| LDpred2 | f | **0.673 [0.644, 0.703]** | 1,385,476/1,386,717 | 0.715 [0.708, 0.722] | 1,354,421/1,386,717 |
|  | r | 0.660 [0.631, 0.690] | 1,377,127/1,378,360 | 0.707 [0.700, 0.714] | 1,346,216/1,378,360 |
|  | s | **0.673 [0.644, 0.702]** | 1,378,226/1,379,459 | 0.712 [0.705, 0.719] | 1,347,305/1,379,459 |
| PRS-CSx | f | 0.672 [0.643, 0.701] | 765,275/766,037 | 0.705 [0.697, 0.712] | 749,451/766,037 |
|  | r | **0.662 [0.633, 0.692]** | 760,636/761,394 | 0.697 [0.690, 0.705] | 744,885/761,394 |
|  | s | 0.669 [0.639, 0.699] | 761,231/761,989 | 0.701 [0.694, 0.708] | 745,477/761,989 |
| PRSice-2 | f | 0.636 [0.605, 0.666] | 1,668/1,668 | 0.653 [0.645, 0.660] | 1,636/1,668 |
|  | r | 0.610 [0.579, 0.641] | 24,476/24,481 | 0.664 [0.657, 0.672] | 24,013/24,481 |
|  | s | 0.637 [0.607, 0.668] | 1,626/1,626 | 0.649 [0.641, 0.657] | 1,594/1,626 |
| Nalls PGS | f | 0.640 [0.610, 0.670] | 1,801/1,805 | 0.663 [0.655, 0.670] | 1,730/1,805 |

AUC: area under the receiver operating characteristic curve [95% confidence interval]. f/ r/ s: PGS construction
based on all genome-wide variants (f: full model) / excluding variants on PD-related genes (r: removed-genes
model) / excluding variants on PD-related genes except for *SNCA* (s: *SNCA* model). #SNPs LuxPark/CouragePD:
number of SNPs from the model available in the LuxPark/CouragePD data / total number of SNPs as in Table 2.
The best-performing tool for f, r and s is highlighted in bold.

**Figure 3:**
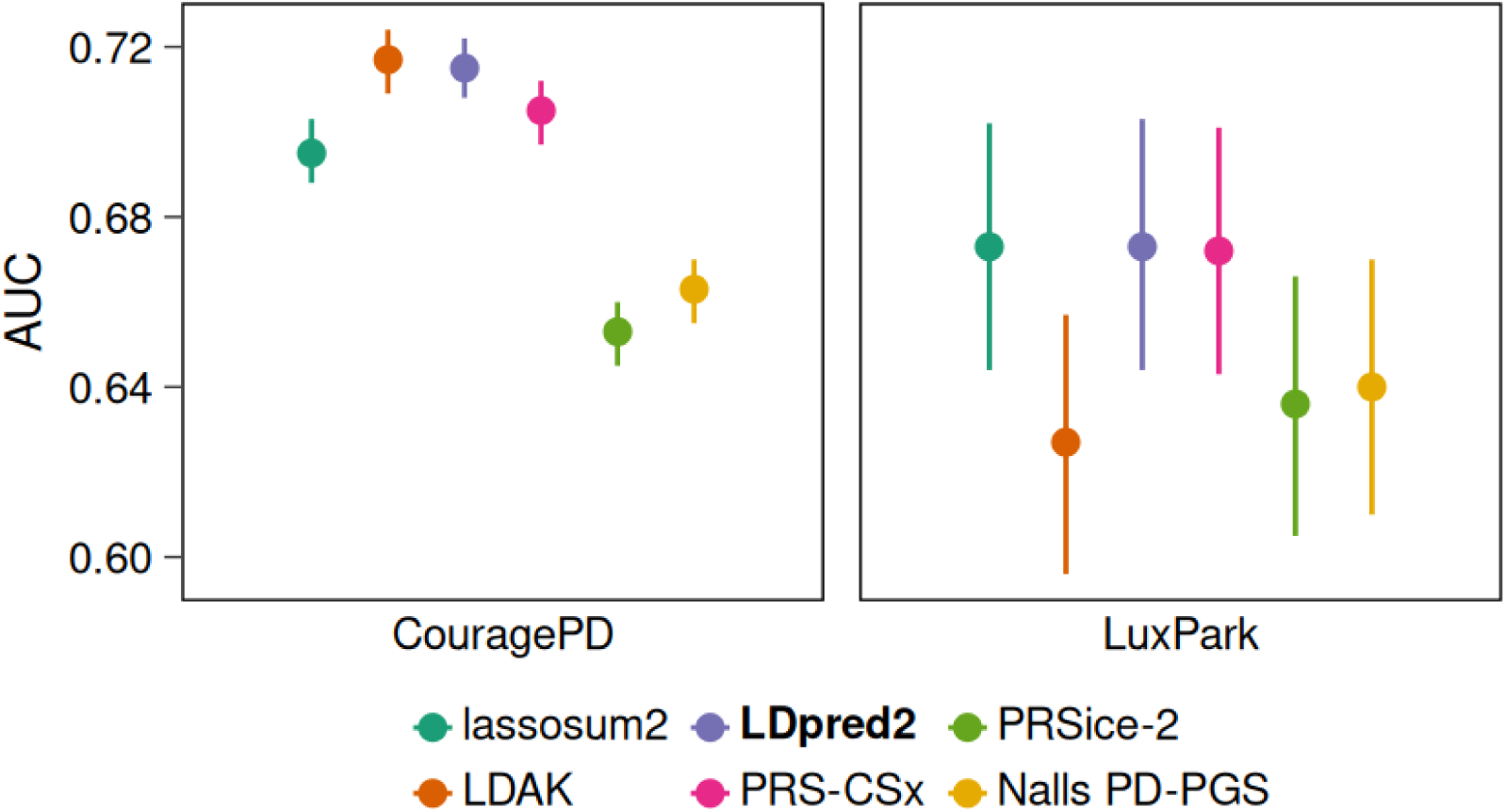
Comparison of performance of constructed iPD-PGSs on external validation datasets. Summary of model performance across the two external validation datasets CouragePD and LuxPark for idiopathic Parkinson’s disease (iPD), using the area under the receiver operating characteristic curve (AUC) as measure. The error bars indicate the 95% confidence interval. Different colors correspond to the selected tools for polygenic score (PGS) generation with the established PGS by Nalls et al.^13^ as comparison. The overall best-performing and selected final tool (LDpred2) is highlighted in bold.

The correlation between PGS values generated by different tools varied considerably (Fig. 4). For example, while a high correlation between the PGSs of LDpred2 with those of lassosum2 and PRS-CSx was observed (Spearman correlation coefficient r of 0.91 and 0.94, respectively, p < 0.001), the PGS values of PRSice-2 and LDAK showed only moderate correlations with the other PGS values (r between 0.51 and 0.80, p < 0.001).

**Figure 4:**
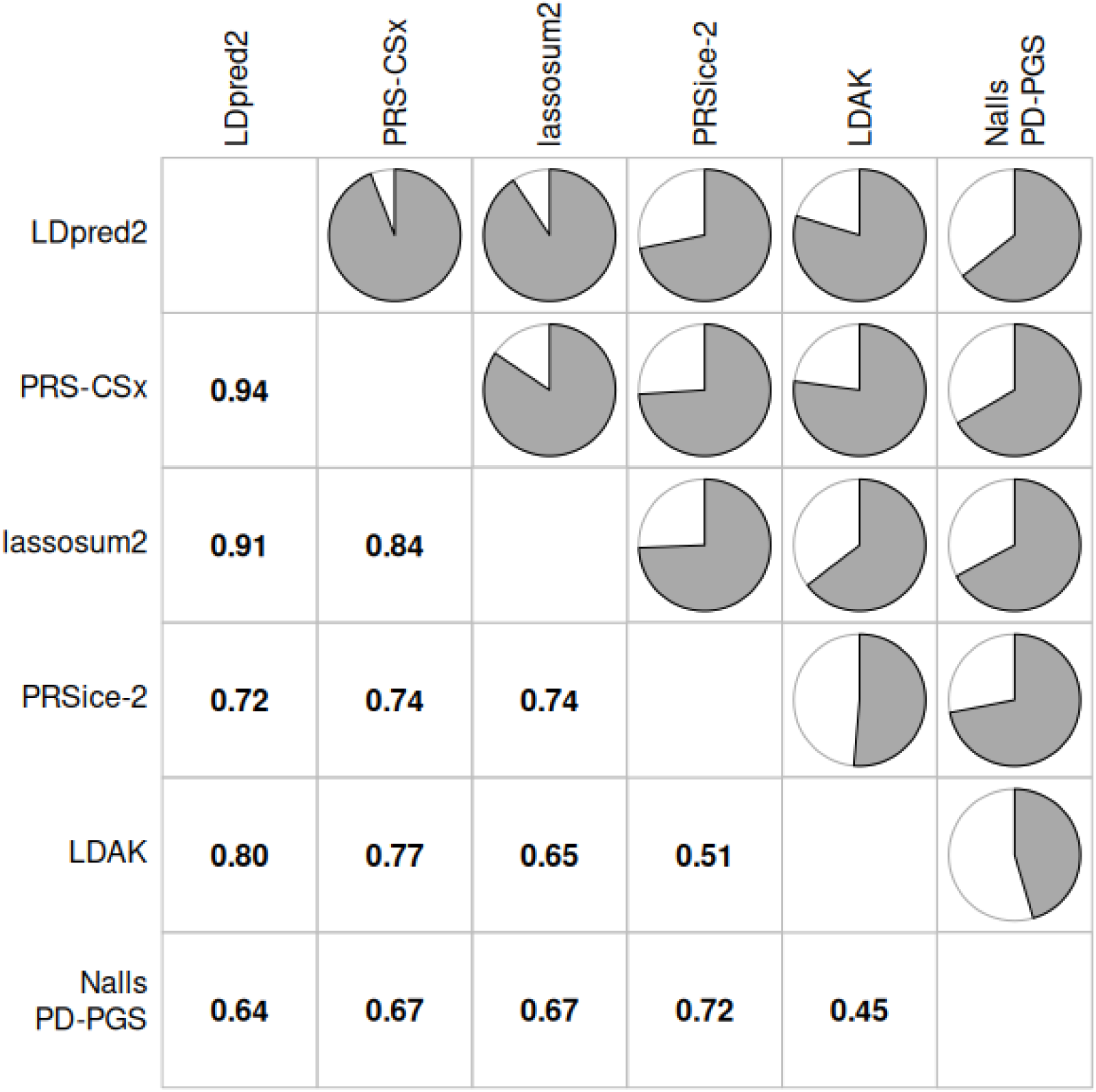
Correlation between PGS values based on different tools. Given are Spearman correlation coefficients calculated for our late-onset idiopathic Parkinson’s disease (iPD) dataset. The established polygenic score (PGS) by Nalls et al.^13^ is shown for comparison.

### Performance of best iPD-PGS

The iPD-PGS generated by LDpred2 demonstrated exceptionally high predictive performance on the ProtectMove late onset iPD and the two external datasets, showing robust results across all three datasets. Therefore, this iPD-PGS served as the basis for our subsequent evaluations in this study (Table 2). Fig. 5A shows the density curves of this PGS for patients and healthy individuals in our whole ProtectMove late-onset iPD dataset. The capability of the PGS to distinguish between patients and healthy individuals was further investigated by dividing the late-onset iPD dataset into deciles of PGS values. We then calculated odds ratios (ORs) by comparing individuals with the lowest PGS values (i.e., in the first decile) with those with higher PGS values (i.e., in the second to tenth deciles) (Fig. 5B). ORs varied between 1.463 (second decile, 95%-CI: [1.051, 2.046]) and 9.342 (tenth decile, 95%-CI: [6.973, 12.664]). The PGS also showed an association with AAO (Fig. 5C). An AUC of 0.698 (95%-CI: [0.637, 0.756]) was achieved when differentiating patients of the first and tenth decile of AAO (AAO<49 and AAO>75) based on the PGS. The correlation between PGS and AAO was -0.178 (95%-CI: [-0.225, -0.129], p < 0.001) and -0.167 (95%-CI: [-0.215, -0.118], p < 0.001) for the Pearson and Spearman correlation, respectively.

**Figure 5:**
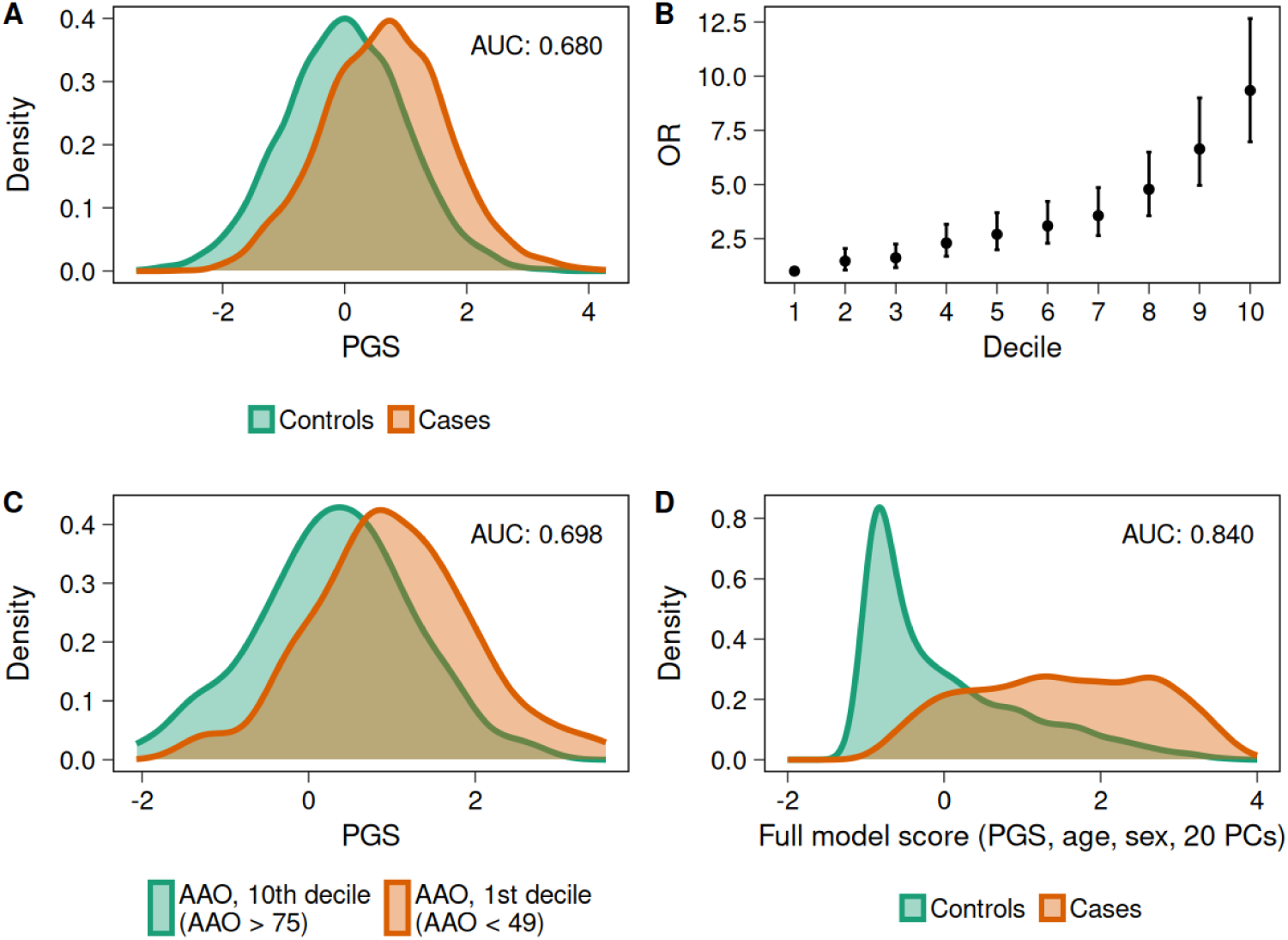
Evaluation of the best-performing iPD-PGS. The best-performing iPD-PGS (LDpred2) was applied to the whole late-onset idiopathic Parkinson’s disease (iPD) dataset. **A:** Density curves of the standardized polygenic score (PGS) for patients and healthy individuals. The discriminative ability of the PGS for these groups is measured by the area under the receiver operating characteristic curve (AUC). **B:** Odds ratio (OR) of case-control status for each PGS decile, compared to the first PGS decile. The error bars indicate the 95% confidence interval. **C:** Density curves of the standardized PGS for cases within the highest and lowest age-at-onset (AAO) decile. **D:** Density curves of the full logistic regression model (influence variables PGS, AAS, sex and the first 20 principal components) for cases and controls.

The discriminative performance of our derived PGS and combined with other covariables is displayed in Table 4. We investigated age (AAS), sex, and the first 20 principal components (PCs). As expected, age alone showed a high discriminative potential (R^2^=0.302, AUC=0.787), whereas the association of either sex or the PCs with PD was lower (sex: R^2^=0.019, PCs: R^2^=0.047, AUC=0.614). Including all three influencing factors (age, sex, PCs) improved model performance (R^2^=0.340, AUC=0.808). Notably, adding our PGS on top of these three factors further enhanced this performance, and a high AUC of 0.840 was achieved (95%-CI: [0.830, 0.851], R^2^=0.407, Fig. 5D), underscoring the strong predictive potential of the combined model (Supplementary Figure 6).

**Table 4:** Performance of the best-performing PGS (LDpred2) combined with covariables on the whole ProtectMove late-onset iPD dataset

| Variables in the model | AUC/counts | R<sup>2</sup> | P-value |
| --- | --- | --- | --- |
| PGS | 0.680 [0.665, 0.695] | 0.119 | 2.989×10<sup>-101</sup> |
| AAS | 0.787 [0.775, 0.799] | 0.302 | 2.474×10<sup>-193</sup> |
| Sex (m/f) | controls: 2046/2172<br>cases: 1058/672 | 0.019 | 1.031×10<sup>-18</sup> |
| PCs (20) | 0.614 [0.598, 0.630] | 0.047 | 1.547×10<sup>-22</sup> (PC1) |
| AAS + sex + 20 PCAs | 0.808 [0.796, 0.819] | 0.340 | 4.496×10<sup>-182</sup> (age) |
| PGS + AAS + sex + 20 PCs | 0.840 [0.830, 0.851] | 0.407 | 4.657×10<sup>-73</sup> (PGS) |

*Variables in the model:* influence factors in the logistic regression model. *AUC:* area under receiver-operating characteristic curve [95% confidence interval]. *counts:* for sex AUC cannot be calculated.
Therefore, the counts for the two categories in controls and cases are given. *R<sup>2</sup>:* Nagelkerke's R<sup>2</sup> of the corresponding logistic regression. *P-value:* p-value of the influence variable in brackets (for more than one influence variable in the model) of the corresponding logistic regression. *PGS:* best-performing PGS of our study (LDpred2). *AAS:* Age-at-sampling. *sex (m/f):* sex (male/female). *PCs:* principal components.

### Influence of PD-related genes

We next investigated to what extent the predictive potential of our developed PGSs was due to genetic variability in genes related to genetic PD (*LRRK2*, *SNCA*, *VPS35*, *CHCHD2*, *PRKN*, *PINK1*, *PARK7*, *GBA1*). We therefore excluded all variants in these eight genes (33,285 out of 5,623,154 variants) from the set of genome-wide variants and recalculated PGSs with all five PGS tools based on this reduced set of variants. The performance of the developed PGSs was slightly reduced, when the PD-related genes were excluded from the construction process (Table 2, column Whole late-onset iPD dataset, AUC no PD-genes). This trend was consistent across all five tools and was also observed in the two external validation datasets (Table 3). For instance, the AUC of our best-performing PGS decreased marginally from 0.680 to 0.672 in the ProtectMove late-onset iPD dataset. To further elucidate the specific role of *SNCA*, we constructed PGSs using a set of variants where those in the *SNCA* gene were retained (4,926), while variants from the other seven PD genes were excluded (28,359). Interestingly, the resulting PGSs demonstrated an equivalent performance to the PGSs including all variants, again demonstrating robustness over all five tools and in the external datasets LuxPark and CouragePD (Table 2, column Whole late-onset iPD dataset, AUC no PD-genes, with *SNCA*, Table 3). The slight reduction in the predictive performance when variants in the eight PD-related genes were excluded is, thus, due to variants in *SNCA*.

### Application of the newly developed iPD-PGS on genetic PD and early-onset iPD

Next, we assessed the role of polygenic background in the variability of penetrance of PD among variant carriers. Our genetic PD dataset consisted of 436 healthy individuals (non-manifesting carriers) and 335 PD patients, both with one (heterozygous) or two (biallelic) genetic causative or risk variants in one of the eight PD-related genes (Table 1, Supplementary Tables 8 and 9.) Most genetic variants were detected in *GBA1* (n=495) and in *PRKN* (n=168 heterozygous and n=14 biallelic). Based on power calculations, we restricted our analysis to these two groups (see Statistical analysis). We considered only carriers of heterozygous genetic variants for *PRKN* because of the difference in disease susceptibility, clinical presentation, and AAO in the biallelic *PRKN* carrier group^37^ (causality can only be assumed in the latter group).

Our best-performing PGS based on all variants demonstrated increased values for *GBA1* variant carriers (both PD patients and healthy individuals; data not shown). This was due to the more common *GBA1* coding risk variants included in the PGS. Therefore, we applied our PGS without variants in PD-related genes to *GBA1* and *PRKN* variant carriers to determine whether the case/control status (and thus the penetrance) could be predicted. The AUC for manifestation of both *PRKN* and *GBA1* variant carriers based on the PGS was significantly larger than 0.5, indicating that the PGS contributes to the varying penetrance in these two groups (Table 5). For *GBA1,* the discriminative potential was larger than for *PRKN* (*GBA1*: AUC=0.639, 95%-CI=[0.590, 0.687], p < 0.001, *PRKN*: AUC=0.594, 95%-CI=[0.501, 0.687], p < 0.0265) (Supplementary Figure 7). Additionally, we examined the group of early-onset iPD. Interestingly, the AUC for iPD patients with an early AAO was even larger than for the late-onset iPD dataset (AUC=0.725, 95%-CI=[0.665, 0.784]).

**Table 5:**
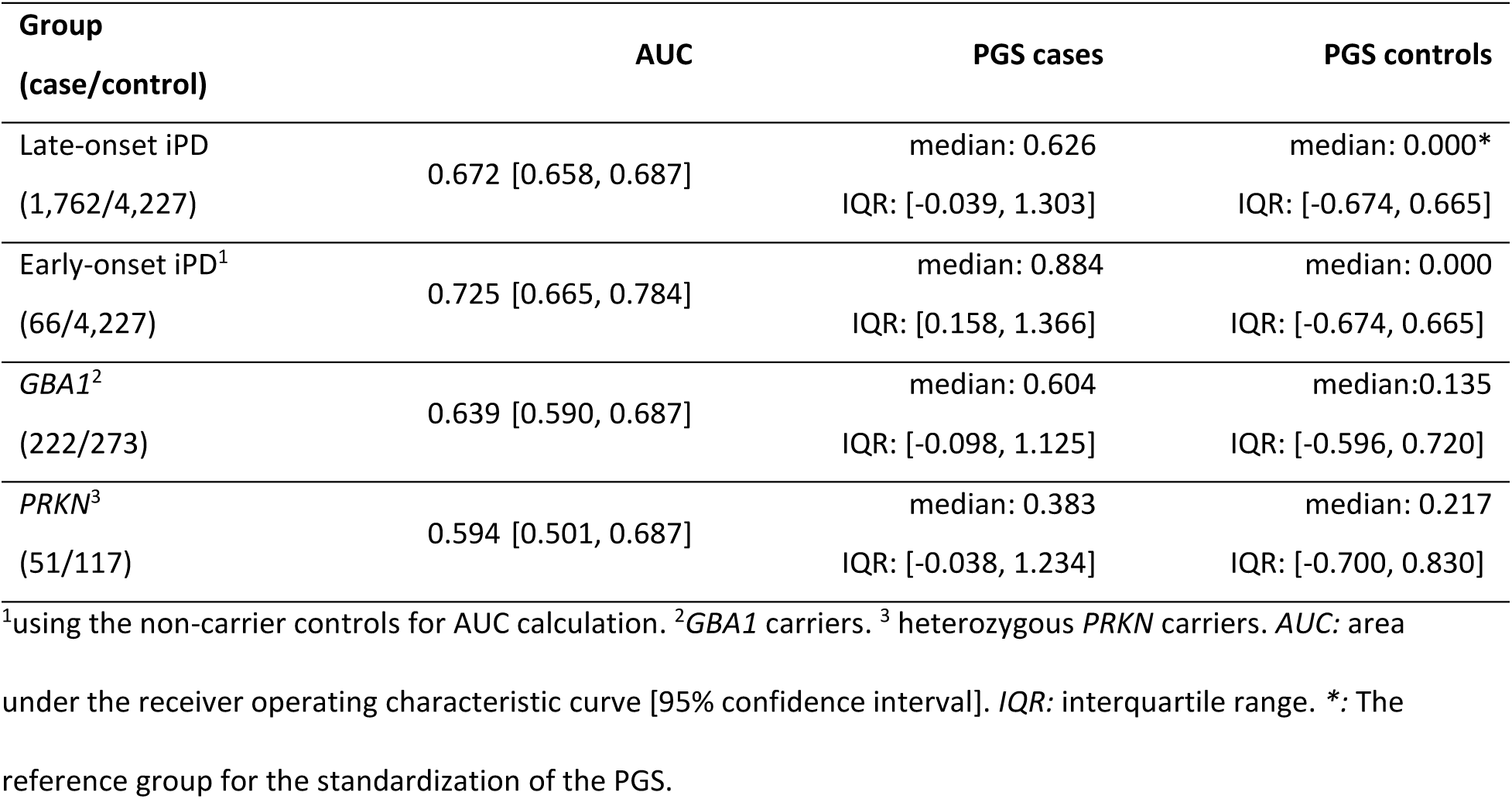
Application of our best-performing PGS without variants on PD-related genes on variant carriers and early-onset iPD patients

| Group<br>(case/control) | AUC | PGS cases | PGS controls |
| --- | --- | --- | --- |
| Late-onset iPD<br>(1,762/4,227) | 0.672 [0.658, 0.687] | median: 0.626<br>IQR: [-0.039, 1.303] | median: 0.000*<br>IQR: [-0.674, 0.665] |
| Early-onset iPD<sup>1</sup><br>(66/4,227) | 0.725 [0.665, 0.784] | median: 0.884<br>IQR: [0.158, 1.366] | median: 0.000<br>IQR: [-0.674, 0.665] |
| *GBA1*<sup>2</sup><br>(222/273) | 0.639 [0.590, 0.687] | median: 0.604<br>IQR: [-0.098, 1.125] | median: 0.135<br>IQR: [-0.596, 0.720] |
| *PRKN*<sup>3</sup><br>(51/117) | 0.594 [0.501, 0.687] | median: 0.383<br>IQR: [-0.038, 1.234] | median: 0.217<br>IQR: [-0.700, 0.830] |

<sup>1</sup>using the non-carrier controls for AUC calculation. <sup>2</sup>*GBA1* carriers. <sup>3</sup> heterozygous *PRKN* carriers. *AUC*: area under the receiver operating characteristic curve [95% confidence interval]. *IQR*: interquartile range. \*: The reference group for the standardization of the PGS.

## Discussion

In this study, we developed a new PGS for iPD that substantially outperformed previously established PGSs^13,38,39^. We achieved this by applying five tools and using three different statistical approaches: Bayesian inference, penalized regression, and clumping-and-thresholding. The performance of PGS models varied substantially depending on the tool used. Among the tools evaluated, LDpred2 consistently demonstrated the best overall performance. It excelled across all five CV sets, the entire training dataset, and both external validation datasets, underlining its robustness across cohorts of the same ancestry. The PRS-CSx’s strong performance is comparable to LDpred2 but is slightly less robust on the CV datasets. The advanced Bayesian framework shared by both tools likely explains their superior performance. It accounts for the LD structure, and thus adjusts the GWAS summary statistic weights and incorporates more markers and information into the PGS. The penalized regression tool lassosum2 delivered robust and reliable results across all datasets but slightly underperformed compared to LDpred2 and PRS-CSx. LDAK (Ridge-SS), another tool based on penalized regression, performed similarly to lassosum2 in most cases but exhibited unique characteristics: it achieved the highest AUC on the CouragePD dataset—outperforming even LDpred2—while delivering the poorest results on the LuxPark dataset. These simpler penalized regression frameworks may offer advantages when computational efficiency is required^34^.

In contrast, PRSice-2 consistently performed the worst among the evaluated methods. This highlights the limitations of more straightforward clumping-and-thresholding methods, which may struggle to capture the entire polygenic architecture of iPD by discarding informative markers^25^. The results of this study exhibit the potential to improve the predictive capability of PGS by advanced statistical methodology.

The number of SNPs included in a PGS model heavily relies on the method used for its development and significantly impacts its performance. Our study demonstrated that models with fewer SNPs, such as those based on the clumping-and-thresholding tool PRSice-2, exhibit the poorest discriminative ability. This finding aligns with previous literature^26^ suggesting that limited SNP inclusion may be insufficient for assessing the complete polygenic architecture of complex diseases. On the other hand, PGS models with fewer SNPs have the advantage of being easier to calculate and potentially simpler to transfer to external datasets that use different SNP arrays. Moreover, a performance drop could occur when applying models with high SNP inclusion on external datasets due to overfitting and differing imputation quality. In our study, the PGS models comprising a high number of variables were robustly validated across two independent datasets. Importantly, this robustness was maintained even without using proxy SNPs reinforcing the integrity of the models.

Furthermore, we tested a sparse version of LDpred2, using only 800,971 SNPs (instead of 1,386,717), which did not yield improved performance on the external validation datasets, suggesting that the inclusion of a broader SNP set is advantageous for predictive accuracy on external data for PD. Genetic variability in eight known PD-related genes does not drive the high predictive potential of our newly derived PGS. Indeed, there was only a slight decrease in the prediction performance when variants on PD-related genes were excluded. Interestingly, there was no decrease at all, when variants on *SNCA* were included while variants on the other seven PD-related genes were excluded. Variants on *SNCA* were thus causal for this small performance drop. *SNCA* is long known to play a major role in monogenic PD, but for our study it seems to have a significant influence in the polygenic inheritance of iPD as well probably due to the risk variants on this gene^40^. Risk variants on *GBA1*, on the other hand, did not contribute to the PGS accuracy.

The performance of our best iPD-PGS varied among different groups. The discriminative ability between PD patients and healthy individuals, measured by AUC, was highest for early AAO patients, followed by non-carriers (late-onset iPD cohort), *GBA1* carriers, and *PRKN* carriers. The iPD-PGS’s ability to distinguish between PD-patients and healthy individuals, even in carriers of *GBA1* and *PRKN* variants, reveals its role in modulating the penetrance of rare causal genetic variants or coding risk variants on *GBA1* by capturing the polygenic background. The lower AUC values observed for *GBA1* and *PRKN* carriers compared to non-carriers highlight the predominant role of rare mutations or specific risk variants in driving disease in these individuals. This study therefore could identify the polygenic background as one factor that explains the varying penetrance in variant carriers of genetic PD, at least for the genes *GBA1* and *PRKN*. This also highlights that genetic and idiopathic PD are not completely separated entities but share the polygenic background as a common risk factor.

A previous study found that a PGS for iPD modulated PD disease risk for *GBA1* carriers^41^. However, the applied PGS consisted only of 90 independent genome-wide significant variants previously identified^13^. In contrast, our PGS was especially derived for iPD with advanced statistical methodology and includes more than one million variants. Another study investigated the interplay of monogenic causal variants and the polygenic etiological component for familial hypercholesterolemia, hereditary breast and ovarian cancer syndrome, and Lynch syndrome. They also found that the polygenic background modified the penetrance of monogenic variants and suggested more research in this important area to bridge the translation to patients^42^.

The superior performance of the iPD-PGS in early-onset individuals suggests that these patients have, on average, an elevated individual score. It is therefore unlikely that the early onset of this group is mainly due to undetected variants in single genes. Instead, they carry a high burden of common risk variants that accelerate disease onset. The polygenic background is therefore particularly influential for these patients, underscoring the predictive potential of iPD-PGSs for individuals with exceptionally high PGS values. This was also targeted in an earlier study by calculating a PGS for early-onset (≤ 50 years) PD patients with and without a monogenic cause. However, the small number of patients (n=90) and the lack of healthy controls render their findings primarily exploratory^43^.

The relevance of PGSs in addition to rare variants in understanding the genetic contribution to PD has high imperative for clinical practice. For non-variant carriers, the iPD-PGS serves as a valuable risk stratification tool. Those individuals with very high PGS values (in the tenth decile) have ninefold increased odds of being a PD patient than persons with very low PGS values (in the first decile).

Although transfer from diagnostic to prognostic ability is limited without longitudinal data, this finding shows the potential for the PGS to serve as a tool in personalized medicine. In addition to differentiating between patients and healthy individuals, the iPD-PGS performs well in discriminating early from late AAO patients. The increased AUC in early-onset individuals underscores the need for targeted research into genetic and environmental factors contributing to early-onset disease. For variant carriers of genetic PD, the PGS provides insights into modifiers within the polygenic background.

It is important to interpret our findings in light of potential limitations. First, we only investigated variant carriers in *GBA1* and heterozygous carriers in *PRKN*. The sample size of variant carriers of other genes was not sufficient for meaningful statistical analyses. Initiatives to increase the number of mutation carriers, such as the Global Parkinson’s Genetics Program (GP2), are ongoing and should result in promising follow-up studies in the future^44^. Second, we defined mutation carriers by single nucleotide variants (SNVs) or copy number variants (CNVs) rated as pathogenic, likely pathogenic, or VUS without differentiation between pathogenicity scoring or variant type (SNV or CNV). This suggests another avenue for future valuable research. A further fundamental limitation is our restriction to European individuals. PGSs are known to be highly ancestry-sensitive^45,46^. Because most GWAS data are available for individuals of European ancestry, this might exacerbate healthcare differences globally^47^. The awareness of the requirement of sufficient genetic data for minorities, diverse and admixed populations, and adequate statistical methodology has been increased in recent years^48^. Several initiatives have also started to address this challenge^49^ for PD, such as GP2^44^.

Developing advanced PGSs for other or multi ancestries or methods for transferring PGSs across ancestries could provide equitable PGS-related medical care in the future.

## Methods

### Study cohorts and variant carrier identification

For this study, we utilized data from nine cohorts (Supplementary Table 10) which were collected in the ProtectMove consortium (https://protect-move.de/). Further information on the cohorts included can be found in the Supplement.

For validation, we had access to two independent datasets. Both datasets were quality controlled to include only individuals of European ancestry. The CouragePD^36^ dataset comprised 10,854 patients and 8,626 healthy individuals. This dataset did not include information on the carrier status of variants on PD-related genes. The LuxPark dataset included 822 patients and 814 healthy individuals, with *LRRK2*, *SNCA*, *VPS35*, *CHCHD2*, *PRKN*, *PINK1*, PARK7 and *GBA1* carriers removed^50,51^.

For the definition of PD-related genes, we restricted our analysis to the confirmed PD-linked genes *LRRK2*, *SNCA*, *VPS35*, *CHCHD2*, *PRKN*, *PINK1*, *PARK7*, and *GBA1*. This selection is based on the published “classical parkinsonism” and “early onset parkinsonism” lists^52^. Additionally, we included carriers of coding risk variants in *GBA1*. Genetic variants (SNVs and CNVs) within these genes were assessed for pathogenicity using the Franklin (https://franklin.genoox.com - Franklin by Genoox), Varsome^53^ and MDSGene^54,55^ assessments. The possible values for pathogenicity are “benign”, “likely benign”, “VUS “, “risk variant” (for *GBA1* only), “likely pathogenic” and “pathogenic”, in increasing order of severity. Individuals were defined as variant carriers of genetic PD if they had at least one allele or one CNV in one of these genes with a pathogenicity rating of at least “VUS”. The final pathogenicity rating for an individual was determined by the highest-ranked mutation present in that individual^50,52,56^. An overview of the identified variant carriers and the pathogenicity of detected variants can be found in Supplementary Tables 8 and 9.

### Ethics statements

The study was conducted according to the guidelines of the Declaration of Helsinki. Informed consent was obtained from all subjects involved in the study.

It was approved by the Ethics Committees of the University of Lübeck, Germany (protocol code 16-039, date of approval 27 September 2019) and the P2N supervisory board, Kiel University, Germany (protocol code 2021-037, date of approval 16 September 2021). The DeNoPa study was approved by the ethics committee of the Physician’s Board Hesse, Germany (approval no. FF89/2008). The Ethics Committee of the Bolzano Health District approved the GESSPARK protocol on 05 June 2008 (30/2008), with an update approved by the Ethics Committee of the Healthcare System of the Autonomous Province of Bolzano-South Tyrol on 11 October 2017. The Ethics Committee of the Bolzano Health District approved the DISP protocol on 25 July 2012 (62/2012), with an update approved by the Ethics Committee of the Healthcare System of the Autonomous Province of Bolzano-South Tyrol on 11 October 2017.

### Genotyping and quality control

For the ProtectMove data, genomic DNA was extracted from peripheral blood leukocytes and genotyped using the Infinium Global Screening Array with Custom Content (GSA; Illumina Inc., San Diego, CA, USA), which targets 645,896 variants. Quality control was performed using PLINK 1.9^57,58^, PLINK 2.0^58,59^, and the R package plinkQC^60^. The following thresholds were applied at the SNP level: minor allele frequency (MAF) ≥ 0.01, SNP call rate ≥ 0.98, and Hardy-Weinberg equilibrium test ≥ 10^-50^ for the software-issued p-value. After this process, a total of 431,738 variants passed quality control and were used for imputation with SHAPEIT2^61^ and IMPUTE2^62^, leveraging the publicly available portion of the HRC reference panel (release 1.1, The European Genome-Phenome Archive, EGAS00001001710). Imputation generated genotyping data for 39,106,911 variants. Variants with MAF < 0.01 or an info score < 0.7 were excluded, leaving a final dataset of 7,804,284 variants for downstream analysis.

We performed quality control for genotyped individuals by removing individuals with increased heterozygosity rate, related individuals and individuals with non-European ancestry (Supplementary Methods). The dataset was then divided into three groups: late-onset iPD (patients and unaffected individuals without variant carriers and with an AAO above 40 years for patients), early-onset iPD (patients without variant carriers and with an AA0 < 40 years) and a genetic PD (affected and not affected variants carriers). See Table 1 for further information on the three groups. The final ProtectMove study population consisted of 6,826 individuals. Subsequent SNP selection resulted in 5,623,154 SNPs for PGS construction.

### PGS construction

#### a) PGS algorithms and GWAS

We constructed PGS models for iPD according to five different tools (PRSice-2^20,21^, LDpred2^25,26^ (inf, auto, grid), lassosum2^34,35^, LDAK^29^ (BayesR-SS, Bolt-SS, Ridge-SS), PRS-CSx^27,28^). While PRSice-2 is based on clumping and thresholding and both lassosum2 and LDAK’s Ridge-SS utilize penalized regression, LDpred2, LDAK’s BayesR-SS, LDAK’s Bolt-SS and PRS-CSx rely mainly on Bayesian approaches. All of the methods use a multitude of parameters in their models, which we varied to obtain the best-performing tool-specific PGS (for more details see Supplementary Methods and Supplementary Table 1). As GWAS summary statistics, we applied the largest GWAS for PD in individuals of European ancestry, including 37,688 cases, 18,618 UK Biobank^63^ proxy-cases, and 1,417,791 controls^13^. The GWAS encompassed a total of 7,784,415 SNPs (6,611,727 without ambiguous SNPs) and were kindly provided by Mike Nalls (with approval by the 23andMe project, agreement University of Kiel SOW #1). The GWAS summary statistics were harmonized to fit the format and effect direction for the ProtectMove late-onset iPD dataset. Ambiguous SNPs, SNPs with a MAF below 0.01 or with a different alternate allele than in the late-onset iPD set were removed. For SNPs with a swapped major and minor allele compared to the late-onset iPD data, the effect size was multiplicated with -1.

#### b) Construction of cross-validation datasets

To identify the best and most robust PGSs for iPD, we performed a five-fold CV. For this, we decomposed the late-onset iPD dataset into the groups “cases with known sex and known AAO”, “controls with known sex and known AAS” and the remainder. We then used the fold() function from the R-package “groupdata2”^64^ on each group to create five equal-sized CV sets. The three groups (cases, controls and remainder) were each divided into five groups of equal size, with each of these five groups being equally distributed according to cohort affiliation, sex and age (mean AAO for cases, mean AAS for controls) and with equally distributed case-control status for the remainder. The resulting CV sets of these three groups were then combined. See Supplementary Table 2 for a more detailed description of the CV sets.

#### c) Construction of PGS models without PD-related genes

To develop a PGS without SNPs from the PD-related genes *LRRK2*, *SNCA*, *VPS35*, *CHCHD2*, *PRKN*, *PINK1*, *PARK7* and *GBA1*, we removed all SNPs within the gene regions with an additional distance of 1,000,000 base-pairs to either side in our genetic data of the late-onset iPD cohort. We also created PGS models in which the SNCA gene was retained while the other seven PD-related genes were removed.

### Statistical analysis

#### a) Identifying the best-performing PGS model

We performed five-fold CV by dividing the dataset into five groups of the same size and the same structure (see above). On each CV step, one set served as the validation set for the PGS models trained on the combined remaining four sets. We carried out the following steps to determine the highest-performing PGS model:

i. For each CV step, various models for each tool (PRSice2, LDpred2, lassosum2, PRS-CSx, LDAK) were calculated depending on the variation of parameters per tool (Supplementary Methods).
ii. Based on the AUC performance, one resulting best model per tool and per training dataset was selected.
iii. The selected models were then applied to the respective validation datasets and mean AUC over the five validation sets calculated to estimate the prediction performance on an independent internal dataset.
iv. We then trained the prediction tool on the whole iPD cohort.
v. The best models of each tool trained on the whole iPD cohort were applied to our two external validation datasets LuxPark and CouragePD.
vi. The best-performing PGS was then identified by the performance on the internal validation datasets of the CV and on the two external validation datasets.

#### b) Statistical measures, models, tests and software

Descriptive measures used in our study were frequencies for categorical variables and median and interquartile range for continuous variables. All statical calculations were performed using the statistics software R^65^, version 4.4.1. All applied statistical tests were two-sided and a significance level of 0.05 was used. All statistical figures have been created using the R package “ggplot2”^66^, unless stated otherwise.

Individual PGS scores were calculated for the training and validation sets using Plink2, based on the selected SNPs and their corresponding weights. The scores were standardized to have a mean of 0 and a standard deviation of 1 among healthy individuals. The receiver operating characteristics (ROC) curve and the AUC, along with 95%-CIs, were computed using the R package “pROC”^67^, the respective p-values were calculated using the R package “verification”^68^. Additionally, we evaluated PGS performance by calculating ORs for individuals within the PGS deciles compared to the first PGS decile for the best-performing model. For the correlation between PGS and AAO, the correlation coefficients and 95%-CIs were calculated using base R (for Pearson) and the *SpearmanRho()* function from the R package “DescTools”^69^ (for Spearman). Furthermore, the densities of individual PGS values for the first versus the tenth AAO decile of the PD-patients were plotted and the corresponding AUC between these AAO groups calculated. To evaluate the role of covariables, we applied six logistic regression models for PD status (case/control) as outcome using R’s *glm()* function, with the following influence variables: (1)-(4) Each of the following variables individually: PGS, age (AAS), sex and the first twenty PCs (PCs included together), (5) all variables except PGS and (6) all aforementioned variables together. Nagelkerke’s R² for each logistic regression model was calculated using the *r2_nagelkerke()* function from the R package “performance”^70^. To estimate the concordance of the PGS values generated by the five different PGS tools, we calculated Spearman correlation coefficients and used the R package “corrplot”^71^ to visualized the results.

For the analysis of variant carriers, we restricted our statistical analysis to the two largest groups, i.e. variants on the genes *GBA1* and *PRKN*. SNVs, as well as CNVs, were considered (see the respective subsection above for the definition of variant carriers). For *PRKN*, only heterozygous carriers were analyzed due to the small number of homozygous carriers and the distinct differences in disease susceptibility, clinical presentation, and AAO between the two groups^37^. Variant carriers with variants on PD-related genes in more than one gene were excluded from this analysis. We applied our best-performing PGS, constructed without variants from PD-related genes, to ensure that the observed performance was attributable to the polygenic component rather than the influence of variants in the gene under investigation, as intended by this analysis. This PGS was also applied to our dataset of early-onset iPD patients (AA0 < 40) and compared against healthy individuals of the late-onset iPD dataset. We calculated the median and interquartile range (IQR) of the PGS for each group, separately for patients and healthy individuals, and the AUC with corresponding 95%-CI.

#### c) Power calculation for analysis of variant carriers

We performed power calculations for variant carriers of different PD-related genes to evaluate which genes were meaningful to analyze. As basis we utilized the Wilcoxon rank-sum test for comparison of PGS values between patients and healthy individuals carrying variants on a specific PD-related gene and assumed a significance level of 0.05. For *GBA1,* 222 patients and 273 healthy individuals were available which resulted in a power of >99% for a medium effect (Cohen’s d=0.5) and of 80% for a small effect (Cohen’s d=0.3). For PRKN we had 51 patients and 117 healthy individuals yielding a power of 82% for a medium effect and of 41% for a small effect. For all other genes, the power was <20% for a medium effect and <10% for a small effect. Power calculations were performed with the statistics software G*Power 3.1.9.4^72^.

## Data availability

The data that support the findings of this study are available on request from the corresponding author. The data are not publicly available due to privacy or ethical restrictions.

## Acknowledgments

We sincerely thank all study participants for their invaluable contributions to this research. We also acknowledge the cohorts that provided data and samples for this study and the investigators who facilitated access to these resources. The funding bodies had no role in the study design, data collection, analysis, interpretation, or manuscript preparation.

We thank Mike A. Nalls for providing us with the list of the 1805 SNPs included in their published PRS (together with reference alleles and effect sizes β).

We thank Manon Sendel for consultation on the discussion and review of language and content of the manuscript.

## Author information

Sebastian Sendel and Zied Landoulsi are shared first authors, Amke Caliebe and Christine Klein are shared last authors.

## Contributions

Conceptualization, A.C., C.K., I.R.K., K.L., P.M., S.S.; methodology, A.C., I.R.K., B.-H.L., P.M., S.S.; formal analysis, D.R.B., Z.L., S.S.; interpretation of data, A.C., C.K., I.R.K., B.-H.L., K.L., P.M., S.S.; resources, A.B., D.B., M.B., N.B., A.F., H.H., E.H., A.A.H., U.M.K., G.K., R.K., L.M.L., W.L., K.L., B.M., M.N., P.P.P., J.P., E.S., M.S. (Manu Sharma), M.S. (Meike Steinbach), C.T., T.U., A.W., M.W.; data curation, M.K., B.-H. L., Z.L., K.L., E.-J.V.; writing—original draft preparation, A.C., S.S.; writing—review and editing, A.B., M. B., N.B., A.C., C.K., I.R.K., L.M.L., K.L., S.S., C.T.; visualization, S.S.; supervision, A.C., P.M.; project administration, C.K.; funding acquisition, A.C., C.K. All authors reviewed and approved the final manuscript.

## References

1. Steinmetz, J. D. et al. Global, regional, and national burden of disorders affecting the nervous system, 1990–2021: a systematic analysis for the Global Burden of Disease Study 2021. Lancet Neurol. 23, 344–381 (2024).

2. Ben-Shlomo, Y. et al. The epidemiology of Parkinson’s disease. Lancet Lond. Engl. 403, 283– 292 (2024).

3. Willis, A. W. et al. Incidence of Parkinson disease in North America. NPJ Park. Dis. 8, 170 (2022).

4. Dorsey, E. R., Sherer, T., Okun, M. S. & Bloem, B. R. The Emerging Evidence of the Parkinson Pandemic. J. Park. Dis. 8, S3–S8 (2018).

5. Bloem, B. R., Okun, M. S. & Klein, C. Parkinson’s disease. The Lancet 397, 2284–2303 (2021).

6. Lüth, T. et al. Interaction of Mitochondrial Polygenic Score and Lifestyle Factors in LRRK2 p.Gly2019Ser Parkinsonism. Mov. Disord. Off. J. Mov. Disord. Soc. 38, 1837–1849 (2023).

7. Gabbert, C. et al. The combined effect of lifestyle factors and polygenic scores on age at onset in Parkinson’s disease. Sci. Rep. 14, 14670 (2024).

8. Huang, Y. et al. Risk factors associated with age at onset of Parkinson’s disease in the UK Biobank. Npj Park. Dis. 10, 1–8 (2024).

9. Reynoso, A. et al. Gene–Environment Interactions for Parkinson’s Disease. Ann. Neurol. 95, 677–687 (2024).

10. Goldman, S. M. et al. Concordance for Parkinson’s disease in twins: A 20-year update. Ann. Neurol. 85, 600–605 (2019).

11. Wray, N. R. et al. Research Review: Polygenic methods and their application to psychiatric traits. J. Child Psychol. Psychiatry 55, 1068–1087 (2014).

12. Lewis, C. M. & Vassos, E. Polygenic risk scores: from research tools to clinical instruments. Genome Med. 12, 44 (2020).

13. Nalls, M. A. et al. Identification of novel risk loci, causal insights, and heritable risk for Parkinson’s disease: a meta-analysis of genome-wide association studies. Lancet Neurol. 18, 1091– 1102 (2019).

14. Verma, A. et al. Diversity and scale: Genetic architecture of 2068 traits in the VA Million Veteran Program. Science 385, eadj1182 (2024).

15. Park, K. W. et al. Ethnicity- and sex-specific genome wide association study on Parkinson’s disease. NPJ Park. Dis. 9, 141 (2023).

16. Pan, H. et al. Genome-wide association study using whole-genome sequencing identifies risk loci for Parkinson’s disease in Chinese population. Npj Park. Dis. 9, 1–11 (2023).

17. Foo, J. N. et al. Identification of Risk Loci for Parkinson Disease in Asians and Comparison of Risk Between Asians and Europeans: A Genome-Wide Association Study. JAMA Neurol. 77, 746–754 (2020).

18. Kim, J. J. et al. Multi-ancestry genome-wide association meta-analysis of Parkinson’s disease. Nat. Genet. 56, 27–36 (2024).

19. Koch, S. et al. Validity and Prognostic Value of a Polygenic Risk Score for Parkinson’s Disease. Genes 12, 1859 (2021).

20. Euesden, J., Lewis, C. M. & O’Reilly, P. F. PRSice: Polygenic Risk Score software. Bioinformatics 31, 1466–1468 (2015).

21. Choi, S. W. & O’Reilly, P. F. PRSice-2: Polygenic Risk Score software for biobank-scale data. GigaScience 8, giz082 (2019).

22. Lambert, S. A. et al. Enhancing the Polygenic Score Catalog with tools for score calculation and ancestry normalization. Nat. Genet. 56, 1989–1994 (2024).

23. Lambert, S. A. et al. The Polygenic Score Catalog as an open database for reproducibility and systematic evaluation. Nat. Genet. 53, 420–425 (2021).

24. PGS Catalog - Parkinson disease [MONDO:0005180] (Polygenic Trait). https://www.pgscatalog.org/trait/MONDO_0005180/ (2024).

25. Vilhjálmsson, B. J. et al. Modeling Linkage Disequilibrium Increases Accuracy of Polygenic Risk Scores. Am. J. Hum. Genet. 97, 576 (2015).

26. Privé, F., Arbel, J. & Vilhjálmsson, B. J. LDpred2: better, faster, stronger. Bioinformatics 36, 5424–5431 (2021).

27. Ge, T., Chen, C.-Y., Ni, Y., Feng, Y.-C. A. & Smoller, J. W. Polygenic prediction via Bayesian regression and continuous shrinkage priors. Nat. Commun. 10, 1776 (2019).

28. Ruan, Y. et al. Improving Polygenic Prediction in Ancestrally Diverse Populations. Nat. Genet. 54, 573 (2022).

29. Zhang, Q., Privé, F., Vilhjálmsson, B. & Speed, D. Improved genetic prediction of complex traits from individual-level data or summary statistics. Nat. Commun. 12, 4192 (2021).

30. Westenberger, A., Brüggemann, N. & Klein, C. Genetics of Parkinson’s Disease: From Causes to Treatment. Cold Spring Harb. Perspect. Med. a041774 (2024) doi:10.1101/cshperspect.a041774.

31. Cook, L. et al. Parkinson’s disease variant detection and disclosure: PD GENEration, a North American study. Brain J. Neurol. 147, 2668–2679 (2024).

32. Polymeropoulos, M. H. et al. Mutation in the alpha-synuclein gene identified in families with Parkinson’s disease. Science 276, 2045–2047 (1997).

33. Klein, C., Hattori, N. & Marras, C. MDSGene: Closing Data Gaps in Genotype-Phenotype Correlations of Monogenic Parkinson’s Disease. J. Park. Dis. 8, S25–S30.

34. Mak, T. S. H., Porsch, R. M., Choi, S. W., Zhou, X. & Sham, P. C. Polygenic scores via penalized regression on summary statistics. Genet. Epidemiol. 41, 469–480 (2017).

35. Privé, F., Vilhjálmsson, B. J. & Mak, T. S. H. lassosum2: an updated version complementing LDpred2. 2021.03.29.437510 Preprint at 10.1101/2021.03.29.437510 (2021).

36. Grover, S. et al. Genome-wide Association and Meta-analysis of Age at Onset in Parkinson Disease: Evidence From the COURAGE-PD Consortium. Neurology 99, e698–e710 (2022).

37. Klein, C., Lohmann-Hedrich, K., Rogaeva, E., Schlossmacher, M. G. & Lang, A. E. Deciphering the role of heterozygous mutations in genes associated with parkinsonism. Lancet Neurol. 6, 652–662 (2007).

38. Bobbili, D. R., Banda, P., Krüger, R. & May, P. Excess of singleton loss-of-function variants in Parkinson’s disease contributes to genetic risk. J. Med. Genet. 57, 617–623 (2020).

39. Chairta, P. P. et al. Prediction of Parkinson’s Disease Risk Based on Genetic Profile and Established Risk Factors. Genes 12, 1278 (2021).

40. Campêlo, C. L. das C. & Silva, R. H. Genetic Variants in SNCA and the Risk of Sporadic Parkinson’s Disease and Clinical Outcomes: A Review. Park. Dis. 2017, 4318416 (2017).

41. Blauwendraat, C., Nalls, M. A. & Singleton, A. B. The genetic architecture of Parkinson’s disease. Lancet Neurol. 19, 170–178 (2020).

42. Fahed, A. C. et al. Polygenic background modifies penetrance of monogenic variants for tier 1 genomic conditions. Nat. Commun. 11, 3635 (2020).

43. Kukkle, P. L. et al. Genome-Wide Polygenic Score Predicts Large Number of High Risk Individuals in Monogenic Undiagnosed Young Onset Parkinson’s Disease Patients from India. *Adv*. Biol. 6, e2101326 (2022).

44. Global Parkinson’s Genetics Program. GP2: The Global Parkinson’s Genetics Program. Mov. Disord. Off. J. Mov. Disord. Soc. 36, 842–851 (2021).

45. Schultz, L. M. et al. Stability of polygenic scores across discovery genome-wide association studies. Hum. Genet. Genomics Adv. 3, 100091 (2022).

46. Ding, Y. et al. Polygenic scoring accuracy varies across the genetic ancestry continuum. Nature 618, 774–781 (2023).

47. Martin, A. R. et al. Current clinical use of polygenic scores will risk exacerbating health disparities. Nat. Genet. 51, 584–591 (2019).

48. Lim, S.-Y. et al. Uncovering the genetic basis of Parkinson’s disease globally: from discoveries to the clinic. Lancet Neurol. 23, 1267–1280 (2024).

49. All of Us Research Program Genomics Investigators. Genomic data in the All of Us Research Program. Nature 627, 340–346 (2024).

50. Landoulsi, Z. et al. Genetic landscape of Parkinson’s disease and related diseases in Luxembourg. Front. Aging Neurosci. 15, (2023).

51. Pachchek, S. et al. Accurate long-read sequencing identified GBA1 as major risk factor in the Luxembourgish Parkinson’s study. Npj Park. Dis. 9, 1–14 (2023).

52. Lange, L. M. et al. Nomenclature of Genetic Movement Disorders: Recommendations of the International Parkinson and Movement Disorder Society Task Force – An Update. Mov. Disord. 37, 905–935 (2022).

53. Kopanos, C. et al. VarSome: the human genomic variant search engine. Bioinformatics 35, 1978–1980 (2019).

54. Kasten, M. et al. Genotype-Phenotype Relations for the Parkinson’s Disease Genes Parkin, PINK1, DJ1: MDSGene Systematic Review. Mov. Disord. Off. J. Mov. Disord. Soc. 33, 730–741 (2018).

55. Trinh, J. et al. Genotype-phenotype relations for the Parkinson’s disease genes SNCA, LRRK2, VPS35: MDSGene systematic review. Mov. Disord. Off. J. Mov. Disord. Soc. 33, 1857–1870 (2018).

56. Landoulsi, Z. et al. Genome-wide association study of copy number variations in Parkinson’s disease. medRxiv 2024.08.21.24311915 (2024) doi:10.1101/2024.08.21.24311915.

57. Purcell, S. & Chang, C. C. PLINK 1.9. www.cog-genomics.org/plink/1.9/.

58. Chang, C. C. et al. Second-generation PLINK: rising to the challenge of larger and richer datasets. GigaScience 4, s13742-015-0047–8 (2015).

59. Purcell, S. & Chang, C. C. PLINK 2.0. www.cog-genomics.org/plink/2.0/.

60. Meyer, H. plinkQC: Genotype Quality Control with ‘PLINK’. (2021).

61. O’Connell, J. et al. A General Approach for Haplotype Phasing across the Full Spectrum of Relatedness. PLOS Genet. 10, e1004234 (2014).

62. Howie, B. N., Donnelly, P. & Marchini, J. A Flexible and Accurate Genotype Imputation Method for the Next Generation of Genome-Wide Association Studies. PLoS Genet. 5, e1000529 (2009).

63. Sudlow, C. et al. UK Biobank: An Open Access Resource for Identifying the Causes of a Wide Range of Complex Diseases of Middle and Old Age. PLoS Med. 12, e1001779 (2015).

64. Olsen, L. R. groupdata2: Creating Groups from Data. R package version 2.0.5 (2024).

65. R Core Team. R: A Language and Environment for Statistical Computing. R Foundation for Statistical Computing, Vienna, Austria (2024).

66. Wickham, H. Ggplot2: Elegant Graphics for Data Analysis. (Springer International Publishing, Cham, 2016). doi:10.1007/978-3-319-24277-4.

67. Robin, X. et al. pROC: an open-source package for R and S+ to analyze and compare ROC curves. BMC Bioinformatics 12, 77 (2011).

68. Gilleland, E. verification: Weather Forecast Verification Utilities. R package version 1.44 (2024).

69. Signorell, A. DescTools: Tools for Descriptive Statistics. R package version 0.99.58 (2024).

70. Lüdecke, D., Ben-Shachar, M. S., Patil, I., Waggoner, P. & Makowski, D. performance: An R Package for Assessment, Comparison and Testing of Statistical Models. J. Open Source Softw. 6, 3139 (2021).

71. Wei, T. & Simko, V. R package ‘corrplot’: Visualization of a Correlation Matrix (Version 0.92). (2024).

72. Faul, F., Erdfelder, E., Lang, A.-G. & Buchner, A. G*Power 3: a flexible statistical power analysis program for the social, behavioral, and biomedical sciences. Behav. Res. Methods 39, 175– 191 (2007).

