## Supplement for "An improved polygenic score for Parkinson’s disease partly explains variable penetrance of genetic Parkinson’s disease"

#### Supplementary methods

##### Cohort information

We utilized data from nine cohorts (Supplementary Table 10) which were collected in the ProtectMove consortium (https://protect-move.de/).

Samples from two PD patient and healthy control cohorts, Kiel PD and Luebeck PD, were recruited mostly locally in Schleswig-Holstein, the northernmost federal state of Germany. The Bolzano PD cohort was drawn from the DISP and GESSPARK studies, both being case-control studies conducted in the Movement Disorders Outpatient Clinic of the Bolzano Hospital (Northern Italy) from 2009 to 2010 (GESSPARK) and 2012 to 2013 (DISP). EPIPARK[1] is a prospective and longitudinal observational single-center study based in Luebeck that focuses on the non-motor symptoms of PD patients. Another prospective and longitudinal observational single-center study is DeNoPa[2] from Kassel in central Germany, aimed at improving the early diagnosis and prognosis of PD. This study includes early untreated PD patients and matched healthy individuals. The PopGen biobank[3,4], maintained by Kiel University, is a central research infrastructure for recruiting case-control cohorts for various diseases. For this study, PopGen contributed 669 PD patients and 3,041 unaffected individuals from the broader Kiel area. Furthermore, we included smaller cohorts (GENEPARK, ROPAD[5], SysMedPD[6]) from Northern Germany that were enriched with mutation carriers because of sampling procedure or family history. Due to the increased risk of unidentified mutation carriers, we therefore only included known carriers from these cohorts.

DISP study

The Dyskinesia, Impulse control disorder and Sleep in Parkinson's disease (DISP, n=223) case-control study was conducted among 124 adults diagnosed with Parkinson's disease visiting the Movement Disorders Outpatient Clinic of the Bolzano Hospital (Italy) and 99 controls with no diagnosis of the disease recruited from non-related co-inhabitants of cases and the general population of the clinic catchment area from 2012 to 2013 to explore and identify biomarkers and environmental factors related to dyskinesia, impulse control, and sleep in Parkinson's disease.

GESSPARK study

The Genetic South Tyrolean Studies of Parkinson's disease (GESSPARK, n=264) case-control study was conducted among 134 adults diagnosed with Parkinson's disease visiting the Movement Disorders Out-patient Clinic of the Bolzano Hospital (Italy) and 130 controls with no diagnosis of the disease recruited from those accompanying cases to their visits and from the general population of the clinic catchment area from 2009 to 2010 to characterize the genetic epidemiology of Parkinson's disease.

Due to quality control, some samples from the DISP and the GESSPARK study were excluded, yielding the final 389 samples for the Bolzano PD cohort. More background on the studies and the available data can be found here: https://doi.org/10.1016/j.nbd.2018.09.016 and https://doi.org/10.1038/s41598-021-99393-8. The samples are housed in the biobank of the Eurac Research Institute for Biomedicine (BRIF6107).

**Quality control for genotyped individuals and subsequent SNP selection**

We calculated the observed heterozygosity rate per individual using the formula $\frac{CT_{OBS}-HOM_{OBS}}{CT_{OBS}}$, where $CT_{OBS}$ and $HOM_{OBS}$ denote the observed counts of heterozygous and homozygous genotypes, both provided by Plink2's --*het* function.[7,8] This was performed on a linkage-disequilibrium (LD) pruned dataset using the --*indep-pairwise* function in Plink2 with a window size of 50 variants, a step size of five variants, and an $r^2$ threshold of 0.2. The LD-pruned dataset retained 186,209 SNPs. We then removed individuals whose observed heterozygosity rate deviated more than 3 standard deviations from the mean over all individuals, as recommended in[9].

Based on the LD-pruned dataset we identified all pairs of individuals with a relatedness above the KING-threshold of $2^{-3.5}$, indicating at least second degree relatedness[10]. From these, we removed individuals in such a way that on the one hand no apparent relatedness was present in the remaining individuals and on the other hand a maximum number of individuals could be kept in our study population. For inclusion, we prioritized variant carriers. Subsequently, we removed individual with non-European ancestry. This was done by performing a principal components analysis (PCA) and calculating the first two PCs from the European cohorts of the 1000 Genomes Project[11], excluding the Finnish cohort. We used the version of the 1000 Genomes Project data included in the Haplotype Reference Consortium (HRC)[12] dataset due to the greater overlap of SNPs with our dataset. We applied k-means clustering to the PC values of the 1000 Genomes Project samples, with k determined by the silhouette method and the gap-method (R-package: "NbClust"[13]), both yielding an optimal k of 3. For each of the three clusters, we defined an ellipse with the mean PC values as the center and four times the standard deviation for PC 1 and 2 as radii (Supplementary Fig. 1A). Then, PC scores according to the 1000 Genomes Project PCA were computed for our ProtectMove dataset and projected onto the 1000 Genomes Project PCs as recommended by the Plink2 project. Individuals from our dataset with PC values outside these ellipses were classified as population outliers and removed (Supplementary Fig. 1B).

Finally we additionally removed those samples identified as outliers based on all relevant PCs according to a PCA on our samples, following the procedure described by Florian Privé[14,15] using the R-package "bigsnpr"[16]. Before applying this procedure, we examined the scree plot for 20 PCs and identified the first five PCs as relevant (Supplementary Fig. 2). We calculated the probabilistic set distance based on the PCs (bigutilsr::prob_dist[17], k = 5 (default, number of nearest neighbors)) and the statistic of outlierness, defined as the ratio of the self-distance to the square root of the distance to the five nearest neighbours (Supplementary Fig. 3). A threshold of 0.167 was chosen, effectively removing the outlier clusters from PC 3 and 4 (Supplementary Fig. 4).

The final study population consisted of 6,826 individuals (see Table 1, Supplementary Tables 8-10 and Fig. 1 for more details). Supplementary Fig. 5 depicts the pairwise PC plots coloured by cohort affiliation after quality control.

The non-imputed genetic dataset before the LD-pruning step was then restricted to this final study population. We repeated the quality control for SNPs, removing those with a minor allele frequency (MAF) below 1%, a call rate missingness exceeding 2%, or above the p-value threshold of $10^{-50}$ for the Hardy-Weinberg-equilibrium[18,19], resulting in 423,107 non-imputed SNPs. The imputed data was also restricted to the study population, and the same quality control as for the non-imputed SNPs was applied, leaving

5,633,017 SNPs. Additionally, SNPs with an INFO-Score below 0.7 were further removed, yielding 5,623,154 SNPs. To avoid issues with some PGS generation tools, we used the *snp_fastImputeSimple()* function from the "bigsnpr" R package to impute missing calls.

**PGS construction tools**

The five PGS construction tools which we applied in this study are listed in Supplementary Table 1.

**PRSice-2** is a software developed by Euesden et al. (PRSice[7,20]), and S Choi et al. (PRSice-2[8]). It is distributed as a C/C++ executable combined with an R script. PRSice-2 utilizes the clumping-and-thresholding approach based on the p-values of SNPs from a GWAS. In the clumping phase, first the SNP with the smallest p-value is selected as the index SNP, and a window size (in kilo base-pairs (kbp)) is placed around it on both sides. Within this window, the correlation of the index SNP with each other SNP is calculated (if no LD reference is used) or checked against the LD structure from a provided reference dataset. SNPs with a correlation exceeding the specified $r^2$ threshold are removed. After that, the SNP with the second smallest p-value is chosen as the new index SNP, and the process is repeated. During the thresholding phase, a series of p-value thresholds are adopted. Only those SNPs selected by the clumping step with a p-value below the threshold are retained for the final model. PRSice-2 then compares the models for all p-value thresholds and selects the best performing one. For our analysis, we employed a grid search over various parameter values to determine the optimal settings for window size and $r^2$ threshold. Specifically, we evaluated window sizes of 250 kbp, 500 kbp, 750 kbp, and 1000 kbp, and $r^2$ thresholds of 0.3, 0.35, 0.4, 0.45, 0.5, 0.55, 0.6, 0.65, 0.7, 0.75, and 0.8. The step size for p-value thresholding was fixed at $5\times10^{-9}$. Additionally, we used three different sources to estimate the LD structure: the European cohorts from the 1000 Genomes Project Phase 3 panel[11], consisting of 503 individuals and 5,598,652 SNPs, the HRC dataset[12] of European ancestry from the Wellcome Sanger Institute, which includes 15,895 individuals and 5,304,658 SNPs, and the study data itself.

The **LDpred** tool, originally introduced by Vilhjálmsson et al.[21] and enhanced by Privé et al.[22] as LDpred2, is implemented in the R package bigsnpr. LDpred2 employs a Bayesian inference approach that adjusts the effect sizes of genetic variants, considering LD between them. It uses prior assumptions about the distribution of effect sizes, specifically a Gaussian mixture. The priors are updated using individual genetic data and GWAS data to obtain posterior effect sizes. Key parameters in LDpred2 are the fraction of causal variants (p) and the heritability of the trait ($h^2$). The LDpred2 framework includes three models: The grid model explores a grid over the parameters p and $h^2$ to find the optimal combination. The $h^2$ parameter is initially estimated from the individual genetic data and weighted by the values 0.7, 1.0, or 1.4. The p parameter is varied across the values: 0.0001, 0.00018, 0.00032, 0.00056, 0.001, 0.0018, 0.0032, 0.0056, 0.01, 0.018, 0.032, 0.056, 0.1, 0.18, 0.32, 0.56, and 1.0. LDpred2 also generates a sparse version for each resulting model where some SNP effect sizes are set to zero, if the respective SNP's probability to be causal is below p. The inf model assumes that all SNPs are causal (p=1), eliminating the need for a grid search over p and $h^2$. The auto model automatically estimates the parameters using an iterative procedure, dynamically updating p and $h^2$. Here, the p parameter iterates through the sequence: 0.0001, 0.00013, 0.00018, 0.00024, 0.00032, 0.00043, 0.00058, 0.00078, 0.001, 0.0014, 0.0019, 0.0025, 0.0034, 0.0046, 0.0061, 0.0082, 0.011, 0.015, 0.02, 0.027, 0.036, 0.048, 0.064, 0.086, 0.12, 0.15, 0.21, 0.28, 0.37, 0.5. Following Privé's recommendations[23], after some initial testing we restricted our analysis to the 1,444,196 variants from the HapMap3+ dataset[24], and this preselection yielded the best results and reduced computation time in our calculations. Since not all variants from the HapMap3+ dataset were available in our data, we created an additional dataset by replacing missing variants with proxy SNPs. We used the HRC dataset to find proxy SNPs in our data that are in LD with HapMap3+ variants ($r^2 \geq 0.9$). The LD structure is incorporated into LDpred via an LD matrix, which can be derived from the individual genetic study data or obtained from external reference datasets. We utilized both approaches, using the LD matrix from the HapMap3+ dataset as an external reference.

In addition to the LDpred2 tool, the R package bigsnpr also includes an implementation of the lassosum algorithm, known as **lassosum2**. Originally introduced by Mak[25] in 2017, the lassosum algorithm is based on penalized regression. The key parameters for lassosum2 are lambda and delta. In penalized regression, lambda controls the L1-regularization of the effect sizes from the GWAS, while delta controls the L2-regularization. We explored a range of values for these parameters, running lambda over a grid of 25 values between 0.000566 and 0.0486, and delta at 0.001, 0.01, 0.1, and 1.

Each combination of lambda and delta generates a PGS model with a proportion of the initial SNPs removed, which is referred to as sparsity.

The **PRS-CSx** tool, introduced by Ruan et al.[26], extends the PRS-CS algorithm developed by Ge et al.[27] to accommodate ancestrally diverse populations. This algorithm was implemented by the authors as a Python script. Like LDpred2, PRS-CSx employs Bayesian inference but is distinct from LDpred2 through the use of different priors. The key parameters for PRS-CSx are a, b, and phi. Parameters a and b define the gamma-gamma prior, determining the distribution of the local, SNP-specific shrinkage parameters psi_i. We used the default values of a=1 and b=0.5 because these yielded the best results in pretesting. The parameter phi controls global shrinkage, reflecting the sparsity of the overall model, and is derived from the individual genetic data. For LD reference panels[28], we utilized data from the 1000 Genomes Project and the UK Biobank[29] (1,287,077 SNPs each). Given that our dataset comprised individuals of European ancestry, we exclusively used cohorts of European descent for this.

Furthermore, we utilized the **LDAK**[30] tool, a comprehensive framework that integrates multiple statistical models and supports various heritability models. For our study, we employed version ldak5.2 of the LDAK software package. Following the authors' recommendations, we selected the BLD_LDAK model for estimating heritability. This model computes SNP heritability using 66 parameters that consider MAF, LD levels, and functional annotations such as coding region locations. These are provided by the BLD-LDAK annotation files recommended by Doug Speed[31], which are also based on the 1000 Genomes Project Phase 3 panel. We utilized our study data to generate the heritability matrix and the SNP correlation matrix. For the prediction algorithm, we selected between Ridge-SS, Bolt-SS and BayesR-SS. The other algorithms like Lasso-SS and Elastic-SS showed decidedly worse performance in pretesting. LDAK automatically selects optimal parameters through cross-validation, using a 90%/10% training/validation split and evaluating 11 different models to determine the best configuration.

#### References


1. Kasten, M. *et al.* Cohort Profile: A population-based cohort to study non-motor symptoms in parkinsonism (EPIPARK). *Int. J. Epidemiol.* **42**, 128–128k (2013).

2. Mollenhauer, B. *et al.* Nonmotor and diagnostic findings in subjects with de novo Parkinson disease of the DeNoPa cohort. *Neurology* **81**, 1226–1234 (2013).
3. Lieb, W. *et al.* Linking pre-existing biorepositories for medical research: the PopGen 2.0 Network. *J. Community Genet.* **10**, 523–530 (2019).
4. Krawczak, M. *et al.* PopGen: Population-Based Recruitment of Patients and Controls for the Analysis of Complex Genotype-Phenotype Relationships. *Community Genet.* **9**, 55–61 (2006).
5. Westenberger, A. *et al.* Relevance of genetic testing in the gene-targeted trial era: the Rostock Parkinson's disease study. *Brain J. Neurol.* **147**, 2652–2667 (2024).
6. Balck, A. *et al.* The role of dopaminergic medication, lipid, and endocannabinoid pathway alterations in idiopathic and PRKN/PINK1-mediated Parkinson's disease – a large-scale targeted metabolomics study. 2024.03.01.24303613 Preprint at https://doi.org/10.1101/2024.03.01.24303613 (2024).
7. Chang, C. C. *et al.* Second-generation PLINK: rising to the challenge of larger and richer datasets. *GigaScience* **4**, s13742-015-0047–8 (2015).
8. Purcell, S. & Chang, C. C. PLINK 2.0. www.cog-genomics.org/plink/2.0/.
9. Anderson, C. A. *et al.* Data quality control in genetic case-control association studies. *Nat. Protoc.* **5**, 1564–1573 (2010).
10. Manichaikul, A. *et al.* Robust relationship inference in genome-wide association studies. *Bioinformatics* **26**, 2867–2873 (2010).
11. Auton, A. *et al.* A global reference for human genetic variation. *Nature* **526**, 68–74 (2015).
12. McCarthy, S. *et al.* A reference panel of 64,976 haplotypes for genotype imputation. *Nat. Genet.* **48**, 1279–1283 (2016).
13. Charrad, M., Ghazzali, N., Boiteau, V. & Niknafs, A. NbClust: An R Package for Determining the Relevant Number of Clusters in a Data Set. *J. Stat. Softw.* **61**, 1–36 (2014).
14. Privé, F., Luu, K., Blum, M. G. B., McGrath, J. J. & Vilhjálmsson, B. J. Efficient toolkit implementing best practices for principal component analysis of population genetic data. *Bioinformatics* **36**, 4449–4457 (2020).

15. Privé, F. Detecting outlier samples in PCA. https://privefl.github.io/blog/detecting-outlier-samples-in-pca/.
16. Privé, F., Aschard, H., Ziyatdinov, A. & Blum, M. G. B. Efficient analysis of large-scale genome-wide data with two R packages: bigstatsr and bigsnpr. *Bioinformatics* **34**, 2781–2787 (2018).
17. Privé, F. bigutilsr: Utility Functions for Large-scale Data. (2021).
18. Wigginton, J. E., Cutler, D. J. & Abecasis, G. R. A Note on Exact Tests of Hardy-Weinberg Equilibrium. *Am. J. Hum. Genet.* **76**, 887–893 (2005).
19. Graffelman, J. & Moreno, V. The mid p-value in exact tests for Hardy-Weinberg equilibrium. *Stat. Appl. Genet. Mol. Biol.* **12**, 433–448 (2013).
20. Purcell, S. & Chang, C. C. PLINK 1.9. www.cog-genomics.org/plink/1.9/.
21. Vilhjálmsson, B. J. *et al.* Modeling Linkage Disequilibrium Increases Accuracy of Polygenic Risk Scores. *Am. J. Hum. Genet.* **97**, 576 (2015).
22. Privé, F., Arbel, J. & Vilhjálmsson, B. J. LDpred2: better, faster, stronger. *Bioinformatics* **36**, 5424–5431 (2021).
23. Privé, F. Polygenic scores and inference using LDpred2. https://privefl.github.io/bigsnpr/articles/LDpred2.html.
24. Privé, F., Albiñana, C., Arbel, J., Pasaniuc, B. & Vilhjálmsson, B. J. Inferring disease architecture and predictive ability with LDpred2-auto. *Am. J. Hum. Genet.* **110**, 2042–2055 (2023).
25. Mak, T. S. H., Porsch, R. M., Choi, S. W., Zhou, X. & Sham, P. C. Polygenic scores via penalized regression on summary statistics. *Genet. Epidemiol.* **41**, 469–480 (2017).
26. Ruan, Y. *et al.* Improving Polygenic Prediction in Ancestrally Diverse Populations. *Nat. Genet.* **54**, 573 (2022).
27. Ge, T., Chen, C.-Y., Ni, Y., Feng, Y.-C. A. & Smoller, J. W. Polygenic prediction via Bayesian regression and continuous shrinkage priors. *Nat. Commun.* **10**, 1776 (2019).
28. Ge, T. PRScsx LD reference panels. https://github.com/getian107/PRScsx (2024).
29. Sudlow, C. *et al.* UK Biobank: An Open Access Resource for Identifying the Causes of a Wide Range of Complex Diseases of Middle and Old Age. *PLoS Med.* **12**, e1001779 (2015).

30. Zhang, Q., Privé, F., Vilhjálmsson, B. & Speed, D. Improved genetic prediction of complex traits from individual-level data or summary statistics. *Nat. Commun.* **12**, 4192 (2021).

31. Speed, D., Holmes, J. & Balding, D. J. Evaluating and improving heritability models using summary statistics. *Nat. Genet.* **52**, 458–462 (2020).

32. Euesden, J., Lewis, C. M. & O'Reilly, P. F. PRSice: Polygenic Risk Score software. *Bioinformatics* **31**, 1466–1468 (2015).

33. Choi, S. W. & O'Reilly, P. F. PRSice-2: Polygenic Risk Score software for biobank-scale data. *GigaScience* **8**, giz082 (2019).

34. Privé, F., Vilhjálmsson, B. J. & Mak, T. S. H. lassosum2: an updated version complementing LDpred2. 2021.03.29.437510 Preprint at https://doi.org/10.1101/2021.03.29.437510 (2021).

#### Supplementary Tables

##### Supplementary Table 1: PGS construction tools used in this study

| Tool | Approach | (Hyper) parameters |
|---|---|---|
| PRSice2[32,33] | clumping + thresholding | LD reference, window size, $r^2$ threshold |
| LDpred2[21,22] | Bayesian inference | method (inf, auto, grid), LD matrix/ SNP selection, sparsity, heritability, proportion of causal variants |
| lassosum2[25,34] | penalized regression | LD matrix/SNP selection, L1 and L2 penalty |
| LDAK[30] | penalized regression | method (Ridge-SS, Bolt-SS, BayesR-SS) |
| PRS-CSx[26,28] | Bayesian inference | LD reference |

(Hyper) *parameters:* (hyper) parameters varied in our PGS development

##### Supplementary Table 2: Characteristics of generated cross-validation datasets

| dataset | N | case/control | Sex (female/male/unknown) | Mean AAS (controls)/AAO (cases) |
|---|---|---|---|---|
| **CV set 1** | 1,194 | 352/842 | 564/622/8 | 58.084/62.249 |
| **CV set 2** | 1,200 | 352/848 | 569/624/7 | 57.925/62.256 |
| **CV set 3** | 1,200 | 353/847 | 570/620/10 | 58.052/62.204 |
| **CV set 4** | 1,202 | 354/848 | 573/617/12 | 58.129/62.468 |
| **CV set 5** | 1,193 | 351/842 | 568/621/4 | 58.170/62.482 |

*N:* number of samples. *CV:* cross-validation

##### Supplementary Table 3: LDpred2 results on cross-validation sets and whole late-onset iPD dataset

| dataset | best ref-data | best $h^2$ | best p | best method | AUC (training) | AUC (validation) |
|---|---|---|---|---|---|---|
| CV 1 | HapMap3+ | 0.0710 | 0.018 | grid | 0.678 [0.661, 0.695] | 0.685 [0.652, 0.717] |
| CV 2 | HapMap3+ | 0.0710 | 0.018 | grid | 0.679 [0.663, 0.696] | 0.681 [0.647, 0.714] |
| CV 3 | HapMap3+ with Proxy | 0.0646 | 0.010 | grid | 0.681 [0.665, 0.698] | 0.677 [0.644, 0.710] |
| CV 4 | HapMap3+ with Proxy | 0.0647 | 0.010 | grid | 0.677 [0.661, 0.694] | 0.691 [0.659, 0.724] |
| CV 5 | HapMap3+ LD-Matrix | 0.0930 | 0.018 | grid | 0.683 [0.667, 0.699] | 0.669 [0.635, 0.702] |
| Whole late-onset iPD | HapMap3+ with Proxy | 0.0649 | 0.010 | grid | 0.680 [0.665, 0.695] | - |

Sparse- or non-sparse-option not mentioned, since the non-sparse-option always performed best. *CV n:* cross-validation run with the nth split as validation set. *best ref-data:* external reference dataset used by the best-performing model. *best $h^2$:* heritability weight parameter of the best-performing model. *best p:* proportion of causal variants parameter of the best-performing model. *best method:* method used (inf, auto or grid) for the best-performing model. *Whole late-onset iPD*: whole late-onset idiopathic Parkinson's disease dataset. *HapMap+ with Proxy:*

HapMap3+ dataset with missing SNPs captured by proxy SNPs from the HRC dataset. *AUC*: area under the receiver operating characteristic curve [95%-confidence interval]

**Supplementary Table 4: lassosum2 results on cross-validation sets and whole late-onset iPD dataset**

| dataset | best ref-data | sparsity | best delta | best lambda | AUC (training) | AUC (validation) |
|---|---|---|---|---|---|---|
| CV 1 | HapMap3+ LD-Matrix | 0.977 | 1 | 0.00485 | 0.662 [0.645, 0.679] | 0.678 [0.646, 0.710] |
| CV 2 | HapMap3+ LD-Matrix | 0.977 | 1 | 0.00485 | 0.668 [0.652, 0.685] | 0.653 [0.618, 0.687] |
| CV 3 | HapMap3+ LD-Matrix | 0.977 | 1 | 0.00485 | 0.670 [0.653, 0.686] | 0.648 [0.614, 0.682] |
| CV 4 | HapMap3+ Proxy | 0.979 | 1 | 0.00480 | 0.660 [0.643, 0.677] | 0.684 [0.651, 0.716] |
| CV 5 | HapMap3+ LD-Matrix | 0.977 | 1 | 0.00485 | 0.665 [0.648, 0.682] | 0.666 [0.633, 0.699] |
| Whole late-onset iPD | HapMap3+ LD-Matrix | 0.977 | 1 | 0.00485 | 0.665 [0.650, 0.680] | - |

*sparsity:* resulting sparsity for the best-performing model. *best delta:* L2-penality parameter for the best-performing model. *best lambda:* L1-penality parameter for the best-performing model. For further details see legend of Supplementary Table 3.

**Supplementary Table 5: PRS-CSx results on cross-validation sets and whole late-onset iPD dataset**

| dataset | best ref-data | AUC (training) | AUC (validation) |
|---|---|---|---|
| CV 1 | UKBB | 0.672 [0.656, 0.689] | 0.690 [0.658, 0.722] |
| CV 2 | UKBB | 0.673 [0.656, 0.689] | 0.683 [0.649, 0.717] |
| CV 3 | 1kg | 0.677 [0.661, 0.694] | 0.677 [0.644, 0.710] |
| CV 4 | UKBB | 0.674 [0.657, 0.690] | 0.685 [0.653, 0.718] |
| CV 5 | 1kg | 0.619 [0.602, 0.637] | 0.607 [0.572, 0.642] |
| Whole late-onset iPD | 1kg | 0.676 [0.662, 0.691] | - |

The default priors (gamma-gamma prior a: 1, gamma-gamma prior b: 0.5) and Markov-chain-Monte-Carlo settings were chosen (iterations: 1000, burn-in: 500, thinning factor: 5). *1kg:* 1000 Genomes Project. *UKBB:* UK Biobank. For further details see legend of Supplementary Table 3.

**Supplementary Table 6: PRSice-2 results on cross-validation sets and whole late-onset iPD dataset**

| dataset | best ref-data | best window size | best $r^2$ threshold | best threshold | AUC (training) | AUC (validation) |
|---|---|---|---|---|---|---|
| CV 1 | none | 750 kbp | 0.3 | 0.000576 | 0.632 [0.614, 0.649] | 0.660 [0.627, 0.692] |
| CV 2 | HRC | 1000 kbp | 0.3 | 0.000800 | 0.643 [0.626, 0.660] | 0.633 [0.599, 0.667] |
| CV 3 | HRC | 500 kbp | 0.3 | 0.000800 | 0.644 [0.627, 0.661] | 0.629 [0.595, 0.663] |
| CV 4 | HRC | 750 kbp | 0.3 | 0.000743 | 0.638 [0.621, 0.655] | 0.651 [0.617, 0.686] |
| CV 5 | 1kg | 750 kbp | 0.3 | 0.000590 | 0.636 [0.619, 0.653] | 0.635 [0.600, 0.669] |
| Whole late-onset iPD | HRC | 500 kbp | 0.3 | 0.000800 | 0.641 [0.626, 0.656] | - |

*best window size:* clumping-window size used for the best-performing model. *best $r^2$ threshold:* threshold for permissible correlation between SNPs during clumping for the best-performing method. *1kg:* 1000 Genomes Project. *HRC:* Haplotype Reference Consortium. *kbp:* kilobase. For further details see legend of Supplementary Table 3.

**Supplementary Table 7: LDAK results on cross-validation sets and whole late-onset iPD dataset**

| dataset | best method | AUC (training) | AUC (validation) |
|---|---|---|---|
| CV 1 | Ridge-SS | 0.659 [0.642, 0.676] | 0.675 [0.642, 0.708] |
| CV 2 | Ridge-SS | 0.663 [0.646, 0.680] | 0.658 [0.624, 0.692] |
| CV 3 | Ridge-SS | 0.664 [0.647, 0.681] | 0.655 [0.620, 0.689] |
| CV 4 | Ridge-SS | 0.661 [0.644, 0.677] | 0.668 [0.634, 0.701] |
| CV 5 | Ridge-SS | 0.664 [0.647, 0.681] | 0.655 [0.620, 0.689] |
| Whole late-onset iPD | Ridge-SS | 0.662 [0.647, 0.677] | 0.657 [0.620, 0.694] |

*best method:* method used (Ridge-SS, Bolt-SS, BayesR-SS) for the best-performing model. For further details see legend of Supplementary Table 3.

#### Supplementary Table 8: Variants in PD-related genes detected in our ProtectMove dataset

| Cohort | VUS | risk variant | likely pathogenic | pathogenic | sum |
|---|---|---|---|---|---|
| **Bolzano PD** | 7 | 10 | 6 | 15 | 38 |
| **DeNoPa** | 6 | 18 | 2 | 2 | 28 |
| **EPIPARK** | 21 | 88 | 13 | 21 | 143 |
| **Kiel_PD** | 2 | 17 | 2 | 4 | 25 |
| **Luebeck PD** | 12 | 73 | 19 | 25 | 129 |
| **Popgen** | 62 | 249 | 30 | 45 | 386 |
| **sccNG** | 2 | 8 | 4 | 8 | 22 |

Included are single-nucleotide variants and copy number variants. *VUS:* variants of unknown significance. *sccNG:* small carrier-enriched cohorts from Northern Germany. The cohorts are: GENEPARK, ROPAD and SysMedPD.

#### Supplementary Table 9: Pathogenicity of variants in PD-related genes in our ProtectMove dataset with regard to genes

| Gene | VUS | risk variant | likely pathogenic | pathogenic | sum |
|---|---|---|---|---|---|
| ***CHCHD2*** | 2 | 0 | 1 | 0 | 3 |
| ***LRRK2*** | 0 | 0 | 1 | 16 | 17 |
| ***SNCA*** | 1 | 0 | 0 | 0 | 1 |
| ***VPS35*** | 19 | 0 | 3 | 0 | 22 |
| ***PARK7*** | 1 | 0 | 3 | 0 | 4 |
| ***PRKN*** | 19 | 0 | 38 | 51 | 108 |
| ***PINK1*** | 10 | 0 | 2 | 7 | 19 |
| ***GBA1*** | 2 | 476 | 31 | 9 | 518 |
| **CNVs** | 77 | 0 | 0 | 44 | 121 |
| **N carriers / (Σ of variants)** | 112 (Σ = 131) | 463 (Σ = 476) | 76 (Σ = 79) | 120 (Σ = 127) | 771 (Σ = 813) |

Included are for the eight single genes only single nucleotide variants. *VUS:* variants of unknown significance. *CNVs:* copy number variants (belonging to one of the eight genes, mostly in *PRKN*). *N (carrier/variants):* number of individuals with the corresponding pathogenicity rating, indicated by the highest pathogenicity of a present mutation. Because an individual can have multiple mutations, the number of individuals differs from the column sum Σ across genes and CNVs.

**Supplementary Table 10: Characteristics of cohorts used in this study (after quality control)**

| Cohort | N total | N cases | N controls | N cases female | N controls female | AAS cases | AAS controls | AAO cases |
|---|---|---|---|---|---|---|---|---|
| **Bolzano PD** | 389 | 197 | 192 | 87 | 120 [14] | 71 [63, 76] | 64 [53, 70] | 64 [56, 70] |
| **DeNoPa** | 241 | 149 | 92 | 51 | 32 | 67 [59, 73] | 67 [62, 70] | 67 [59, 73] |
| **EPIPARK** | 1,264 | 520 | 744 | 200 | 353 | 69 [60, 76] | 67 [61, 71] | 60 [52, 70] |
| **Kiel PD** | 181 | 181 | 0 | 60 | 0 | 68 [61, 77] | - | 59 [49, 68] |
| **Luebeck PD** | 1,019 | 426 | 593 | 152 [1] | 361 | 68 [56.75, 75] | 44 [35, 48] | 60 [51, 67] |
| **Popgen** | 3,710 | 669 | 3,041 | 262 [42] | 1,516 [6] | 70 [65, 77] | 54 [40, 65] | 64 [56, 70] |
| **sccNG** | 22 | 21 | 1 | 6 | 1 | 58 [50.25, 66.75] | 78 | 50 [32, 58] |
| **Total** | 6,826 | 2,163 | 4,663 | 818 [43] | 2,383 [20] | 69 [61, 76] | 55 [42, 67] | 62 [54, 70] |

*N total:* total number of samples. *N cases/controls:* number of cases/controls. *N cases/controls female:* number of female [or unknown sex] cases/controls. *AAS cases/controls:* median [first and third quartile] for age of cases/controls in years. *AAO cases:* median [first and third quartile] for AAO of cases in years. *NA:* not available. *sccNG:* small carrier-enriched cohorts from Northern Germany. the cohorts are: GENEPARK, ROPAD and SysMedPD.

### Supplementary Figures

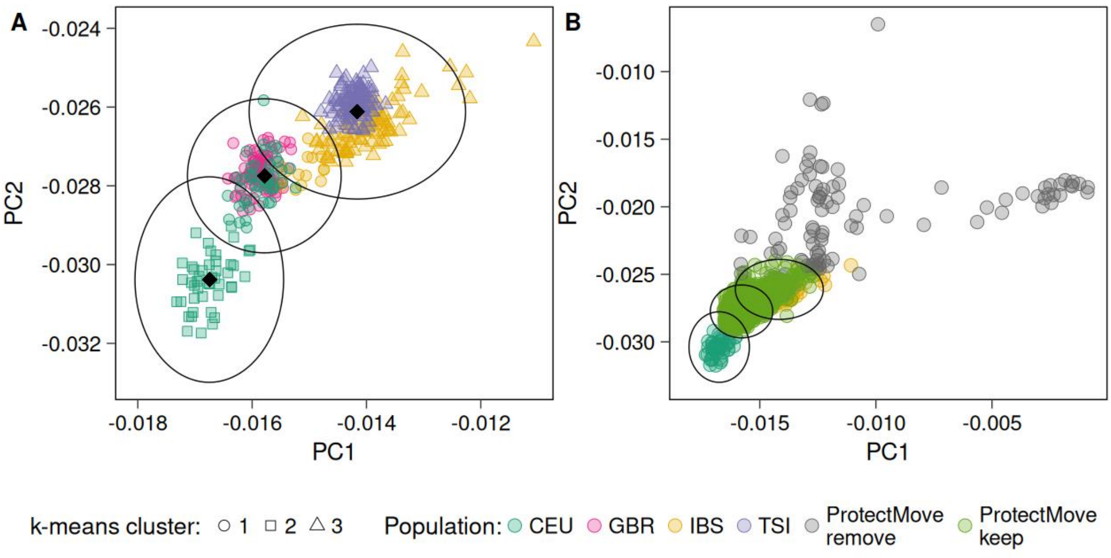


**Supplementary Figure 1: Principal component analysis with 1000 Genomes Project individuals and our study population**

**A:** First two principal components for 1000 Genomes Project individuals, colored by 1000 Genomes Project cohorts and shapes according to the k-means clustering groups.

**B:** First two principal components for 1000 Genomes Project individuals and our ProtectMove data, colored by 1000 Genomes Project cohorts or removal status. Because the ProtectMove data are plotted on top, 1000 Genomes Project cohorts GBR, TSI and IBS are hardly visible here.

*CEU:* Utah residents with Northern and Western European ancestry. *GBR:* British in England and Scotland. *IBS:* Iberian populations in Spain. *TSI:* Toscani in Italy. *PC:* Principal component*. ProtectMove keep/remove:* individuals from our ProtectMove data inside/outside the ellipses that are kept/removed from the dataset.

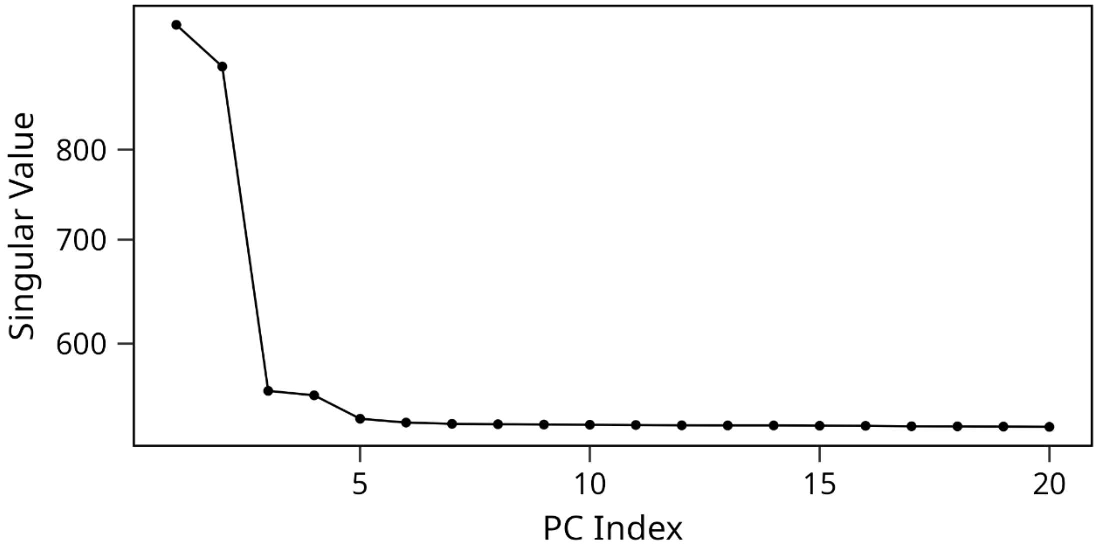


**Supplementary Figure 2: Screeplot across the first 20 principal components**

As a result, we have limited ourselves to five principal components.

*PC Index:* Principal component index

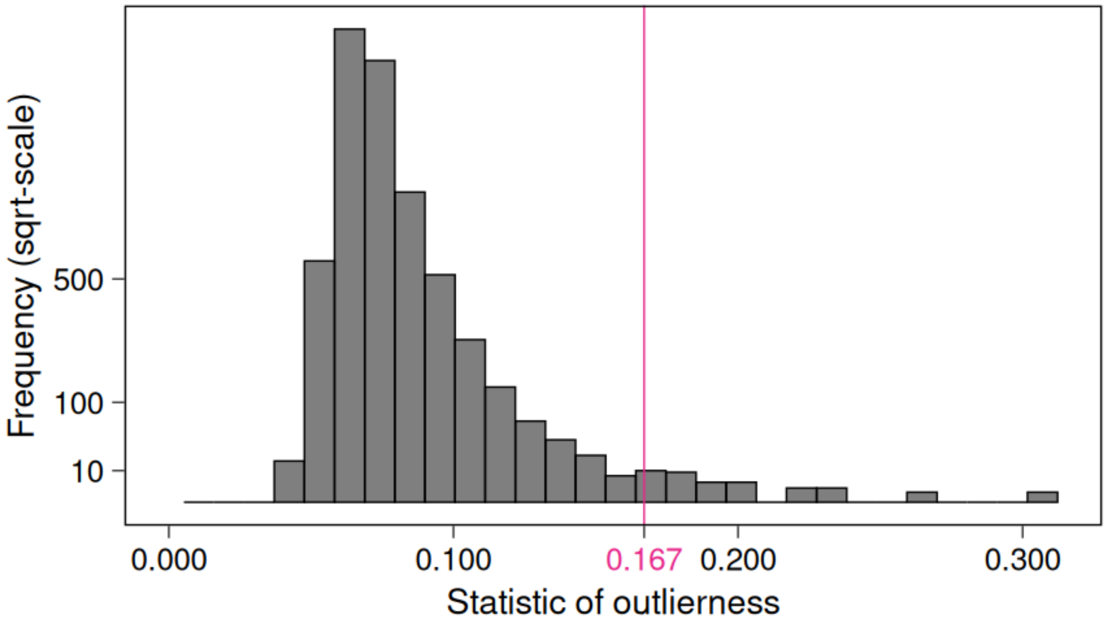


**Supplementary Figure 3: Distribution of the statistic of outlierness for our study population**

0.167 was selected as threshold and any sample above this was discarded from the final dataset.

*sqrt-scale*: square-root-scale of the frequencies on the y axis

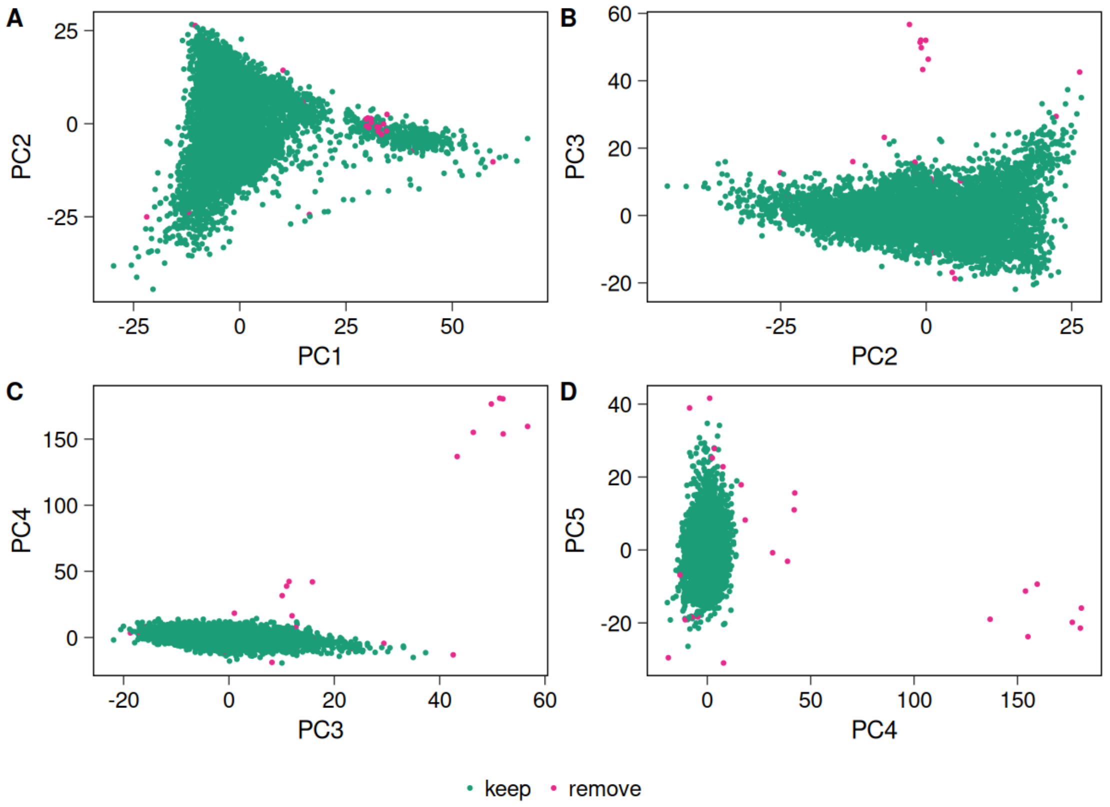


**Supplementary Figure 4: Principal components plots showing the removed outliers**

Samples were removed if their statistic of outlierness was above 0.167.

*PC*: Principal component

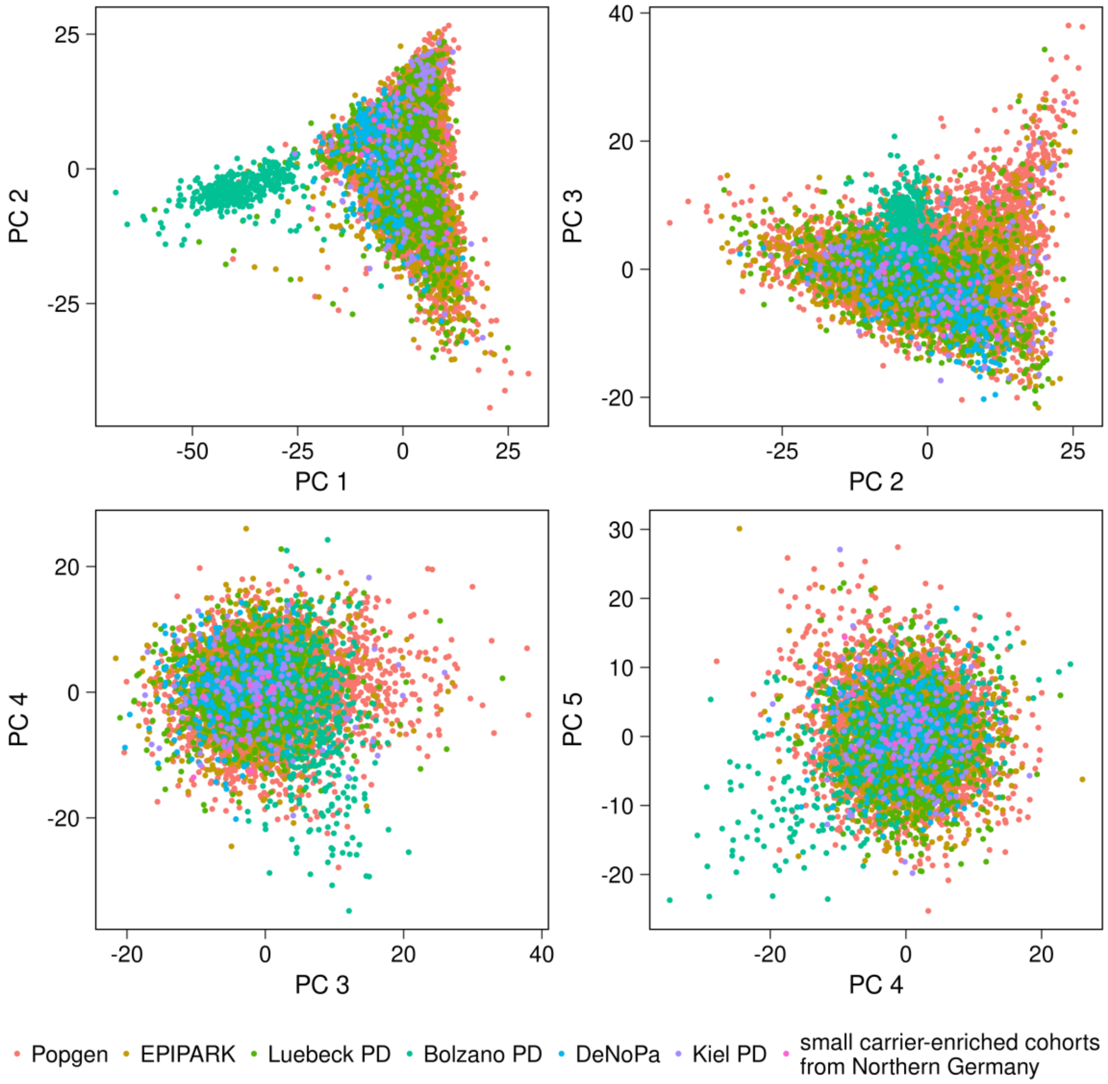


**Supplementary Figure 5: Principal components plots of the final study population colored by cohort**

*PC*: Principal component

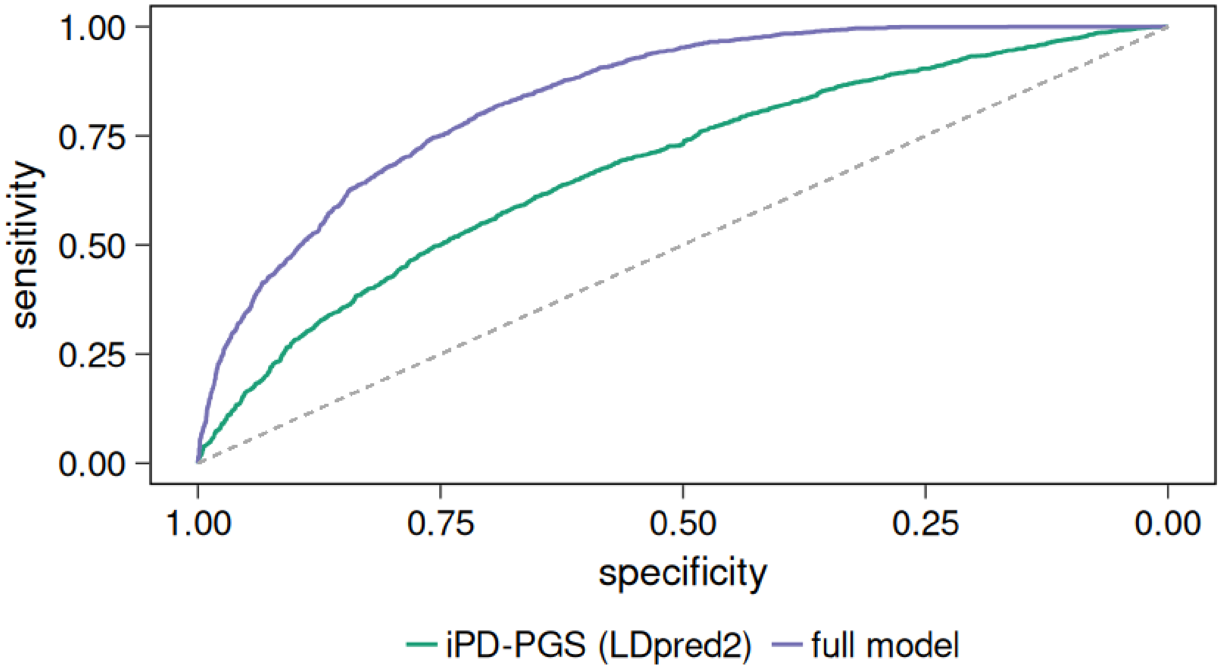


**Supplementary Figure 6:** ROC curve for the best-performing iPD-PGS (LDpred2) and the full model

*ROC:* Receiver operating characteristic. *full model:* Logistic regression model with the iPD-PGS, age-at-sampling, sex and the first 20 principal components as independent variables.

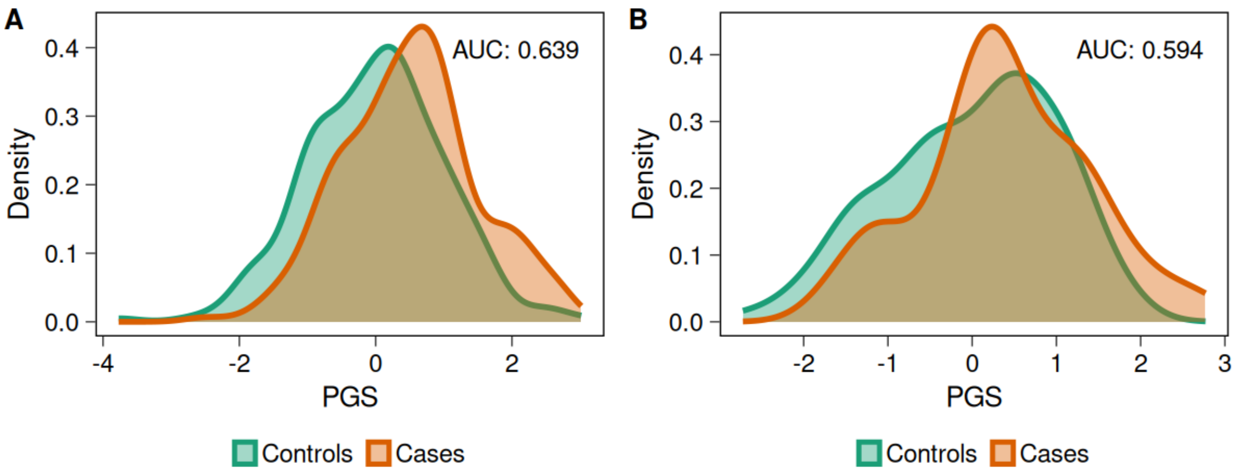


**Supplementary Figure 7:** Density curves for **A:** *GBA1* variant carriers and **B:** heterozygous *PRKN* variant carriers

The best-performing iPD-PGS (LDpred2) without variants in PD-related genes was used.

*AUC:* Area under the receiver operating characteristic curve.
